# Improving ascertainment of suicidal ideation and suicide attempt with natural language processing

**DOI:** 10.1101/2022.02.25.22271532

**Authors:** Cosmin A. Bejan, Michael Ripperger, Drew Wilimitis, Ryan Ahmed, JooEun Kang, Katelyn Robinson, Theodore J. Morley, Douglas M. Ruderfer, Colin G. Walsh

## Abstract

Methods relying on diagnostic codes to identify suicidal ideation and suicide attempt in Electronic Health Records (EHRs) at scale are suboptimal because these phenotypes are heavily under-coded. We propose to improve the ascertainment of suicide phenotypes using natural language processing (NLP). We developed information retrieval methodologies to search over 200 million notes from the Vanderbilt EHR. Suicide query terms were extracted using word2vec. A weakly supervised approach was designed to label cases of suicidal outcomes. The NLP validation of the top 200 retrieved patients showed high performance for suicidal ideation (area under the receiver operator curve [AUROC]: 98.6, 95% confidence interval [CI]: 97.1−99.5) and suicide attempt (AUROC: 97.3, 95% CI: 95.2−98.7). Case extraction produced the best performance when combining NLP and diagnostic codes and when accounting for negated suicide expressions in notes. Overall, we demonstrated that scalable and accurate NLP methods can be developed to identify suicide phenotypes in EHRs to enhance prevention efforts, predictive models, and precision medicine.

## INTRODUCTION

Accurately ascertaining self-injurious thoughts and behaviors from longitudinal clinical data remains a core challenge in prediction, phenotyping, and clinical monitoring, all of which provide clinical utility and support life-saving intervention. Widely used diagnostic classification codes for suicide phenotypes, including suicidal ideation and suicide attempt, are frequently under-coded and under-reported.^1-3^ Prior work has shown the positive predictive value of using diagnostic codes for suicide attempt to be as low as 58.63% in a sample of 5,543 charts, though these estimates range up to near perfect performance.^3,4^ Issues with ascertainment undermine accurate estimates of rates of suicidal outcomes, appropriate resource allocation, quality improvement, and risk assessment.^2,5,6^

Like other behavioral health traits, alternative ascertainment approaches via patient self-report, health information exchange, public health surveillance, and natural language processing (NLP) have been tested to assess improvement from diagnostic codes.^7-11^ The latter, NLP, has been used to improve ascertainment of social determinants of health to augment effective sample size for clinical modeling.^12,13^

Suicidal ideation and suicide attempt share attributes common to phenotypes that lack biomarkers or reliable structured data representation. NLP provides a scalable means of extracting relevant signal to identify such phenotypes using clinical unstructured text. For example, NLP was used to ascertain likelihood of adverse child events or homelessness over time at scale in an electronic health record (EHR).^12^

In this study, we developed and validated NLP methodologies to ascertain 1) suicidal ideation and 2) suicide attempt from clinical notes in a large EHR repository. We compared this NLP approach to diagnostic codes using a gold standard patient cohort obtained through multi-reviewer manual chart validation.

## METHODS

This study presents a scalable NLP approach that receives as input a list of text expressions describing a phenotype of interest (phenotype query), scans all clinical notes from an EHR, and computes a phenotype relevance score for each patient with input text expressions in its notes. The output of this NLP system is a ranked list of patients as potential cases for the phenotype of interest such that the most relevant patients in the list are ranked at the top. The study was approved by the institutional review board at Vanderbilt University Medical Center (VUMC).

### Clinical population

The clinical data used in this study were extracted from Synthetic Derivative, a research-oriented data repository that contains the de-identified version of the VUMC’s EHR.^14^ As of December 2021, this repository stores >200 million notes for >3.4 million patients. Specific data elements extracted from Synthetic Derivative include clinical notes, psychiatric forms, demographics data, and International Classification of Diseases, 9th/10th Revision, Clinical Modification (ICD-9/10-CM) billing codes.

### A data-driven approach to guide the selection of suicide query terms

We relied on a data-driven approach to automatically extract text expressions that describe suicidal ideation and suicide attempt. Similar to our previous work,^12^ we used Google’s word2vec (https://code.google.com/p/word2vec/) to iteratively expand an initial list of 2 relevant seed keywords, *‘suicide’* and *‘suicidal’*. Briefly, we first trained a skip-gram model of word2vec^15^ on 10 million notes randomly sampled from Synthetic Derivative to learn word embeddings for every word in the note collection. The preprocessing of these notes included tokenization, conversion of tokens to lowercase, and exclusion of low-frequency tokens and punctuations. For model configuration, we used a vector dimension of 100, and context window sizes of 5 and 15. Next, we computed the cosine similarity between the seed embeddings and the embeddings of all non-seed words and selected the top ranked words as new seed words and potential candidates for suicide query terms. Finally, we manually analyzed the generated seed list to propose queries for the two suicide phenotypes.

### Retrieval of suicidal ideation and suicide attempt

We implemented an information retrieval model to rank patients by their relevance to each suicide query constructed in the previous step. The system architecture was designed as a vector space model where input queries and patients were represented as multidimensional vectors of words or word expressions. Here, each patient vector was extracted from a meta-document that included all patient notes. The relevance score of a patient to a suicide phenotype was measured as the similarity between the corresponding patient vector and suicide query vector using the standard term frequency-inverse document frequency (TF-IDF) weighted cosine metric. Specifically, for the similarity score between a suicide query and patient *j*, the weight of the query term *i* in the meta-document of the patient *j* was computed as:

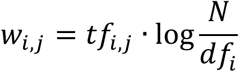

where *tf*_*i,j*_ is the number of occurrences of term *i* in the meta-document of patient *j* (term frequency), *df*_*i*_ is the number of patients whose corresponding meta-documents contain the term *i* (document frequency), and *N* is the total number of patients in the EHR.

For each retrieved patient, we also implemented assertion strategies based on the frequency of negated query terms in patient notes.^16-18^ To assess if negation improves the retrieval of suicidal ideation and suicide attempt, we extracted additional rankings in which each patient has at least one positively asserted query term in patient notes. Thus, these rankings do not contain patients for whom all the query term mentions in their notes are negated.

### Model assessment

Model performance was assessed for both suicidal ideation and suicide attempt on patient sets extracted from three sources of information: 1) top ranked patients extracted by the NLP system, 2) randomly selected patients with ICD10CM codes for self-injurious thoughts and behaviors, and 3) randomly selected patients with psychiatric forms for suicide assessment. Only a limited set of psychiatric forms for suicide assessment was available in Synthetic Derivative because not all structured forms are currently de-identifiable at scale without risking inadvertent re-identification. Each patient was double reviewed by manual analysis (reviewers KR, RA) of the entire patient record and conflicts were resolved by a clinician with expertise in medicine and in chart validation for suicide research (CGW). The inter-reviewer agreement was measured using Cohen’s kappa statistic. Overall, a patient was manually labeled as a case if the corresponding patient notes contain non-zero evidence of suicidal intent. Patients with ICD codes for self-injurious thoughts and behaviors were also required to have supporting information in their notes to be labeled as cases. In situations where a patient denied a suicide attempt, but a clinician documented that an attempt had occurred, the chart reviewers followed the provider’s judgment and assigned a case label.

The evaluation consisted of comparing the patient assessments through manual review with the automatically generated assessments by the NLP system, ICD10CM codes, and psychiatric forms for suicidal ideation and suicide attempt. For the unranked patients, we measured the performance values in terms of precision (P) or positive predictive value (PPV), recall (R), and F1 score (F1). For the ranked patient lists generated by the NLP system, we reported precision-recall curves, precision of top K highest ranked patients (P@K), and area under the precision-recall curve (AUPRC), which was estimated based on the average precision measure.^19^ We employed a bootstrap procedure to compute the 95% confidence intervals (CIs) of the AUPRC estimators using the empirical quantiles of the resampled data generated by 1,000 bootstrap replicates.^20,21^

### A weakly supervised approach to label cases of suicidal ideation and suicide attempt

The main objective of this study was to perform a high-precision extraction of suicidal ideation and suicide attempt cases from all patients extracted by the NLP system. Since we designed the NLP system to rank the most relevant patients for the two suicide phenotypes at the top of each list, we proposed to solve this task by first finding a threshold value, K, for a given target precision, P@K, and then selecting the top K ranked patients from the retrieved list as cases. In our experiments, we extracted K values such that P@K=90% and P@K=80%.

To compute P@K for any K in a ranked list (denoted as *patient*[1..*N*], where *K* ≤ *N*), we designed a weakly supervised approach that assigns a case label to each patient in the list with a specific confidence value or probability (**Algorithm 1**). This approach combines a small set of patients labeled as cases or non-cases with the remainder set of unlabeled patients in the ranked list. We defined the initial labeled set to include all patients from the ranked list that were manually validated or that had psychiatric forms for suicidal ideation and suicide attempt assessment. Based on our evaluation, we assumed each patient from this initial set was labeled as a case or non-case with high confidence (or with a probability *p* = 1). This is specified by the *resultValidation* procedure in **Algorithm 1**.

#### Algorithm 1 A weakly supervised method of case label assignment for a ranked list of patients retrieved by the NLP system

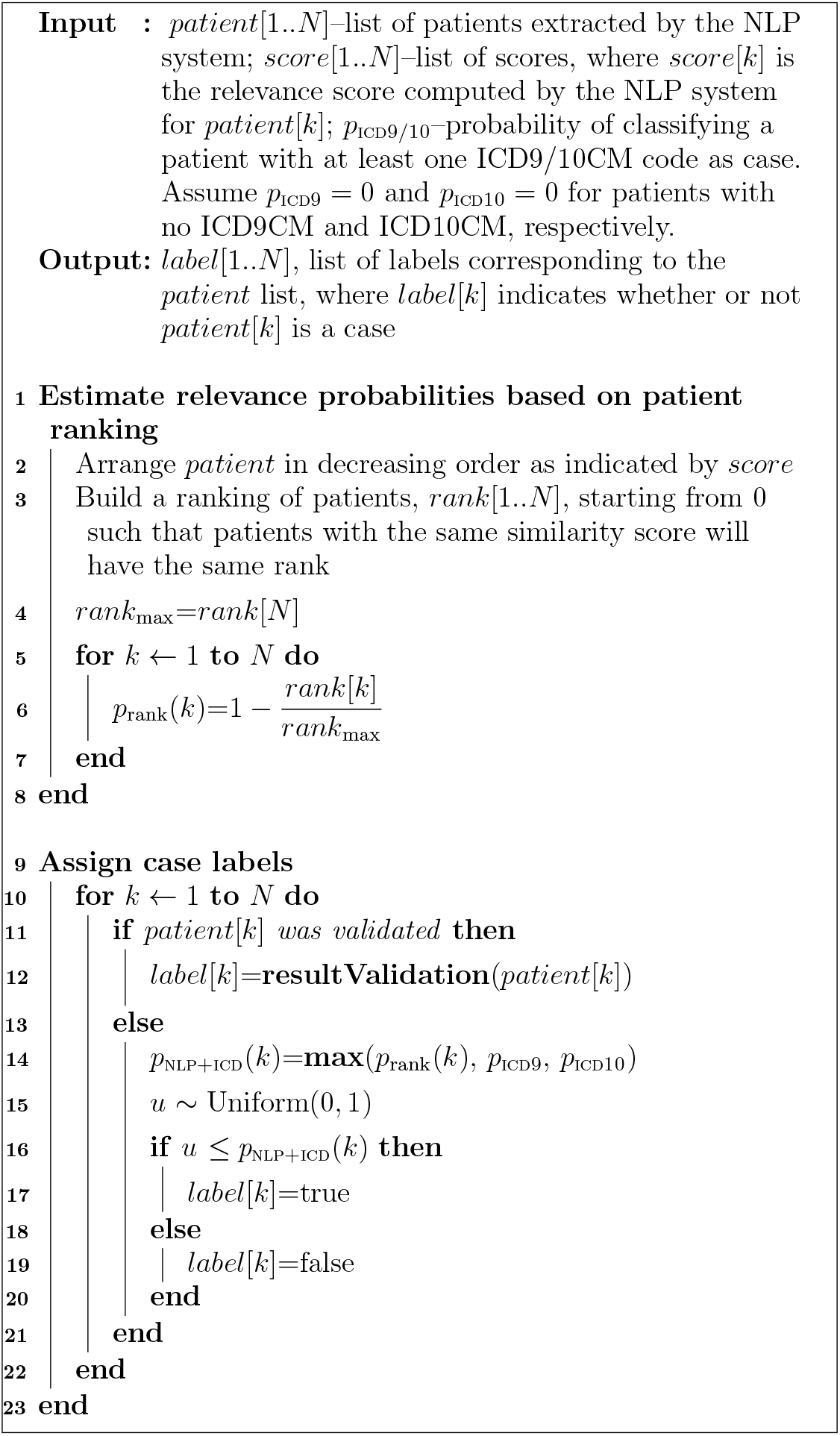

The probability of case assignment for an unlabeled patient was computed according to its rank in the list and availability of relevant ICD codes in its record (**Algorithm** 1, lines 13–21). Specifically, for each patient in the ranked list, we initially computed a relevance probability (denoted as *p*_rank_) that is proportional to the rank position of the patient in the list as described in lines 1–8 in **Algorithm 1**. As observed, *p*_rank_ = 1 for the first patient in the list; then, *p*_rank_ decreases monotonically to 0, which corresponds to the relevance probability of the last patient in the list. Further, based on the evaluation performed in this study and our previous work,^4^ we computed *p*_ICD9_ and *p*_ICD10_ as probabilities of having a suicide phenotype for every patient with at least one relevant ICD10CM and ICD9CM, respectively. We assumed these probabilities to be zero for patients with no ICD codes for self-injurious thoughts and behaviors. When both NLP ranks and ICD codes were considered, we computed the probability of assigning patient *k* to a case label as *p*_NLP+ICD_(*k*) = max(*p*_rank_(*k*), *p*_ICD9_, *p*_ICD10_) as shown by line 14 in **Algorithm 1**. Thus, using this probability and a random variable *u* generated from the standard uniform distribution, the label assignment for patient *k* was performed as indicated by lines 15–20. Additionally, to evaluate the contribution of ICD codes to the selection of suicidal ideation and suicide attempt cases, we implemented a similar weakly supervised approach using only *p*_rank_ probabilities for case assignment. This NLP-based case assignment method was performed by replacing line 14 in **Algorithm 1** with *p*_NLP_(*k*) = *p*_rank_(*k*). Notably, *p*_NLP1ICD_ and *p*_NLP_ could be also set to a minimum value of 0.5 assuming that each patient in the ranked list had at least an equal chance to be randomly assigned to a case. However, this approach will not contribute to the selection of top K cases at P@K=90% or P@K=80% and will result in mainly increasing the number of cases in the bottom half of the ranked patient list where *p*_rank_(*k*) < 0.5. The ICD9CM and ICD10CM codes for self-injurious thoughts and behaviors used in this study are listed in **Tables S1**-**S4**.

## RESULTS

### Suicide query term extraction

The top 50 keywords extracted by word2vec as semantically similar to ‘*suicide*’ and ‘*suicidal*’ under various configurations are listed in **Table S5**. Words from this table including ‘*ideation*’, ‘*self-harm*’, ‘*mutilation*’, and ‘*thoughts*’ were added to the set of seed keywords for suicide, which was further expanded through an iterative approach involving word2vec and manual assessment. Based on this set of seed keywords, we constructed the queries for retrieving the potential cases of suicidal ideation and suicide attempt (**Table S6**). Additionally, during this process, we used words like ‘*thinking*’ and ‘*wanting*’ in suicide expressions to better capture suicidal ideation in notes (e.g., ‘*thinking to kill herself*’, ‘*wanting to end his life*’); similarly, we used words including ‘*attempted*’ and ‘*tried*’ to construct more specific suicide attempt queries (e.g., ‘*attempted to shoot himself*’, ‘*tried to take her own life*’).

### Patient retrieval

Based on the queries identified from the word2vec approach described above (**Table S6**), the NLP system retrieved 187,047 and 52,738 potential cases of suicidal ideation and suicide attempt, respectively (**Table 1**). In these cohorts, only 12.9% (N=24,053) of patients in the suicidal ideation list and 23.5% (N=12,393) in the suicide attempt list had at least one ICD code for self-injurious thoughts and behaviors. Furthermore, the patients with relevant ICD codes were ranked towards the top of the two ranked lists. For example, there were 17,257 (36.9%) of 46,761 patients with relevant ICD codes in the first quartile of the suicidal ideation list compared to 4,210 (9%) of 46,762 patients with relevant ICD codes in the second quartile of the same list. Here, for each list, the first quartile contained the highest ranked patients. Similarly, for the suicide attempt list, the proportion of patients with relevant ICD codes in the first quartile was 53.7% (7,086 of 13,184) compared to 21.4% (2,820 of 13,185) of patients with relevant ICD codes in the second quartile. For these two examples, the proportions of patients with ICD codes for self-injurious thoughts and behaviors in the first and second quartile were significantly different (for both tests, *p* < 2.2 × 10^−16^). An example with the proportions of patients with relevant ICD codes across the two ranked lists of suicidal ideation and suicide attempt is illustrated in **Figure 1**.

**Table 1.** Characteristics of patients retrieved by the NLP system. The extraction of cases and non-cases from psychiatric forms and chart review was restricted to the patients retrieved using NLP. The cases from psychiatric forms have at least one positive field while the non-cases have all the fields negated.

| Characteristic | SI |  | SA |  |
| --- | --- | --- | --- | --- |
|  | N | % | N | % |
| Total patients retrieved | 187,047 |  | 52,738 |  |
| Patients w/ ICD codes | 24,053 | 12.9 | 12,393 | 23.5 |
| Patients w/ 1+ positive mentions | 93,690 | 50.1 | 50,108 | 95.0 |
| **Manual validation** |  |  |  |  |
| Cases | 921 | 0.5 | 682 | 1.3 |
| Non-cases | 79 | 0.04 | 138 | 0.3 |
| **Psychiatric forms** |  |  |  |  |
| Cases | 4,484 | 2.4 | 2,164 | 4.1 |
| Non-cases | 4,308 | 2.3 | 1,380 | 2.6 |

**Figure 1.**
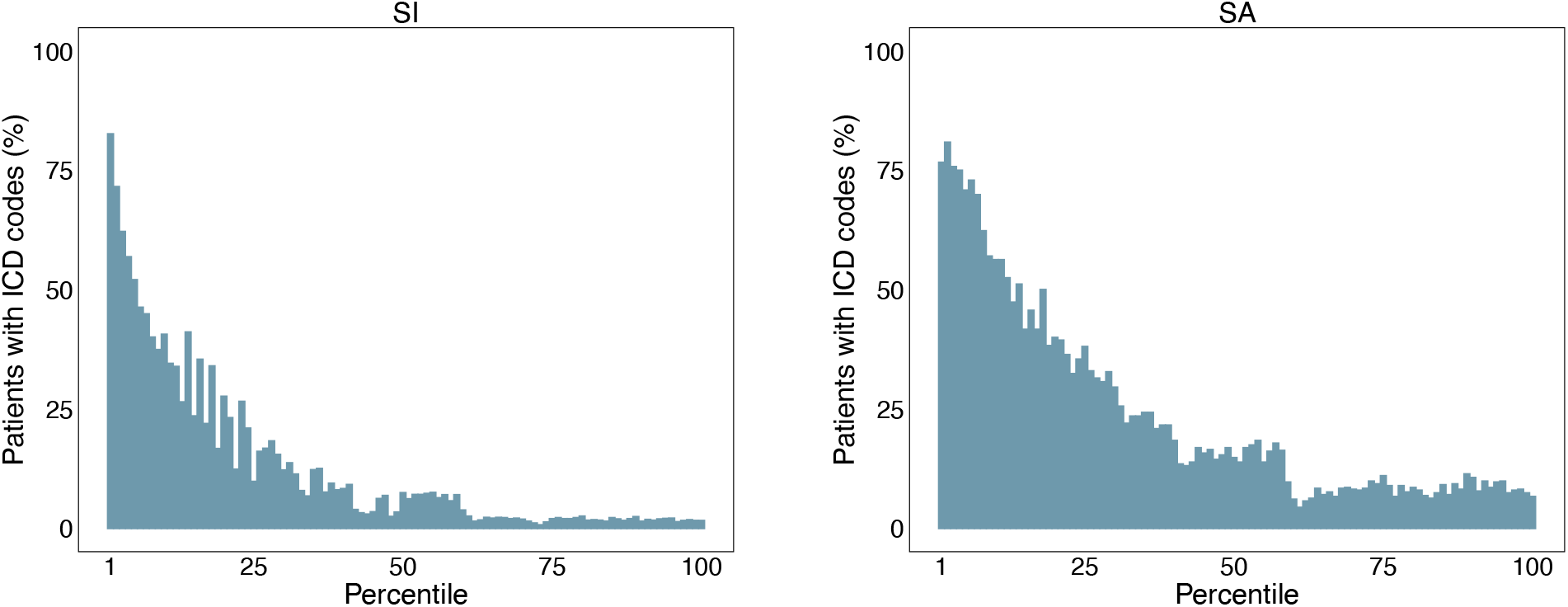
Distribution of patients with ICD codes across the ranked lists of suicidal ideation (SI) and suicide attempt (SA) patients. For each retrieved list, patients were first ordered by their similarity score (or rank position) such that the most relevant ones are ranked at the top of the list. Each list was then split into 100 equal groups (percentiles) with the first and last percentiles representing the highest and lowest ranked patients, respectively. The percent of patients with relevant ICD codes was computed for each percentile.

### NLP-based validation

We manually reviewed and labeled each patient in the top 200 highest ranked patients of suicidal ideation and suicide attempt lists. The manual review of these patients indicated a very high performance of the NLP system (**Table 2A** and **Figure 2**) for both suicidal ideation (P@200: 98.5%; AUPRC: 98.6, 95% CI: 97.1–99.5) and suicide attempt (P@200: 96.5%; AUPRC: 97.3, 95% CI: 95.2–98.7). As expected, the ICD-based evaluation in the top 200 highest ranked patients from these two lists suggests that patients with ICD codes for self-injurious thoughts and behaviors yield a better precision when compared with the patients without any of these codes: 100% vs. 90% for suicidal ideation and 98.7% vs. 90.2% for suicide attempt (**Table 2B**). We also assessed the role of negation in the evaluation of the two suicide phenotypes. However, all the patients in the top 200 highest ranked patients of the suicidal ideation list had at least one positively asserted suicide mention in their notes while only one patient in the top 200 highest ranked patients of the suicide attempt list had all its suicide mentions negated. This patient was manually labeled as non-case; thus, excluding this patient from the top 200 highest ranked patients further improved the precision of extracting suicide attempt cases from 96.5% to 97% (**Tables 2A** and **2B**).

**Table 2.** NLP and ICD10CM validation.

| **A Patient selection** | **Phenotype** | **P@200** | **AUPRC (95% CI)** |
| --- | --- | --- | --- |
| Top 200 patients retrieved by the NLP system | SI | 98.5 | 98.6 (97.1 - 99.5) |
|  | SA | 96.5 | 97.3 (95.2 - 98.7) |

| **B Patient selection** | **Phenotype** | **N** | **P** |
| --- | --- | --- | --- |
| Patients in top 200 w/ relevant ICD codes | SI | 170 | 100 |
|  | SA | 149 | 98.7 |
| Patients in top 200 w/o relevant ICD codes | SI | 30 | 90.0 |
|  | SA | 51 | 90.2 |
| Patients in top 200 w/ 1+ positive mentions | SI | 200 | 98.5 |
|  | SA | 199 | 97.0 |

| **C Patient selection** | **Phenotype** | **N** | **P** |
| --- | --- | --- | --- |
| Patients with ICD10CM codes for suicide | SI | 200 | 96.0 |
|  | SA | 200 | 85.0 |

AUPRC: area under the precision-recall curve, P@K: precision at top K retrieved patients, P: precision, SI: suicidal ideation, SA: suicide attempt

**Figure 2.**
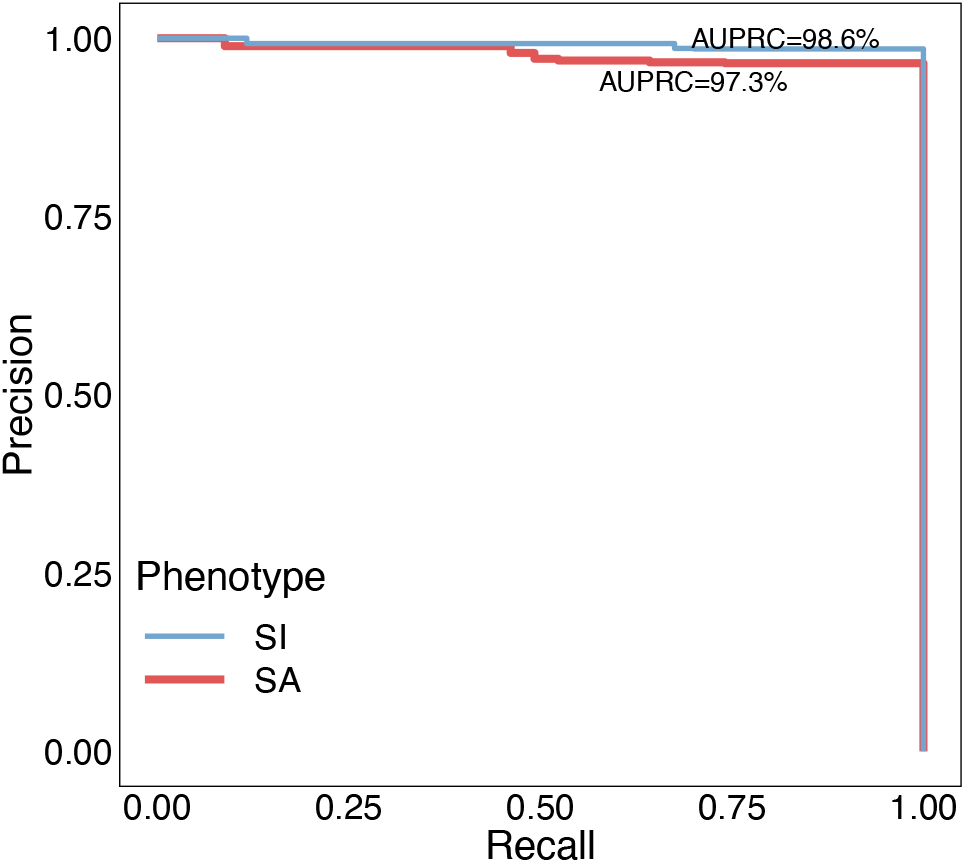
Precision-recall curves for suicidal ideation (SI) and suicide attempt (SA) evaluation of the top 200 highest ranked patients retrieved by the NLP system.

### ICD10CM-based validation

We performed an unbiased evaluation of 200 randomly selected patients with relevant ICD10CM codes for each suicide phenotype (**Table 2C**). The manual review involving the patients selected for ICD10CM and NLP-based validation achieved a substantial interrater agreement for both suicidal ideation (Cohen’s *κ* = 0.72) and suicide attempt (Cohen’s *κ* = 0.8). The ICD10CM-based validation revealed high precision values of 96% and 85% for suicidal ideation and suicide attempt, respectively. Notably, the precision obtained for suicide attempt significantly outperformed the precision of 58.63% we achieved in our previous ICD9CM-based evaluation of this phenotype.^4^

### Evaluation of psychiatric forms for suicide assessment

A random sample of 10 patients with psychiatric forms containing Yes/No information on suicidal ideation and suicide attempt were manually reviewed and all patients were found in perfect agreement with the data encoded in their forms. As a result, the patients with psychiatric forms on the two suicide phenotypes from the NLP-retrieved lists were labeled by the weakly supervised method as case or non-case with high confidence (**Algorithm** 1, lines 11–13).

### Label assignment evaluation for suicidal ideation and suicide attempt

We ran our proposed weakly supervised approach on suicide label assignment leveraging the small set of patients manually labeled as cases or non-cases, the psychiatric forms, the NLP ranks and ICD billing codes. The label assignment over all the patients retrieved by the NLP system enabled us to: 1) assess the overall impact of negation detection in case identification, and 2) evaluate the contribution of ICD codes for the selection of cases. For example, the label assignment methods based on NLP ranks showed an increase in AUPRC for suicidal ideation (from 55.2 to 57.5) and suicide attempt (from 44.5 to 45.2) when only the patients with at least one positively asserted mention of suicide in their notes (instead of all patients) are included in the two lists retrieved by the NLP system (**Table 3**, NLP and All retrieved vs. NLP and w/ 1+positive columns). After excluding the patients with all their suicide mentions negated in notes, in addition to NLP ranks, ICD codes yielded an even more substantial AUPRC increase from 57.5 to 62.2 for suicidal ideation and from 45.2 to 54.1 for suicide attempt (**Table 3**, w/ 1+ positive and NLP vs. w/ 1+ positive and NLP+ICD columns; **Figure 3**, top plots). This trend was also reflected in extracting the top K highest ranked patients from the NLP lists for both P@K=90% and P@K=80%. As a result, higher K values (e.g., for suicidal ideation and P@K=90%, K_NLP_=980 vs. K_NLP+ICD_=1,321) were obtained when negation detection together with NLP ranks and ICD codes were considered for label assignment (**Table 3** and **Figure 3**, bottom plots). To gain a deeper insight into accurately identifying cases, we ran the label assignment algorithm 1,000 times on each ranked list and reported descriptive statistics of the top K highest ranked patients for a target precision, P@K. For instance, leveraging the suicide attempt list of patients with positively asserted mentions of suicide, the label assignment method based on NLP ranks extracted on average 395±38 cases with precision ≥90% whereas the method using both NLP ranks and ICD codes identified on average 527±68 cases at the same level of precision (**Figure 4D** and **Table S5**).

**Table 3.**
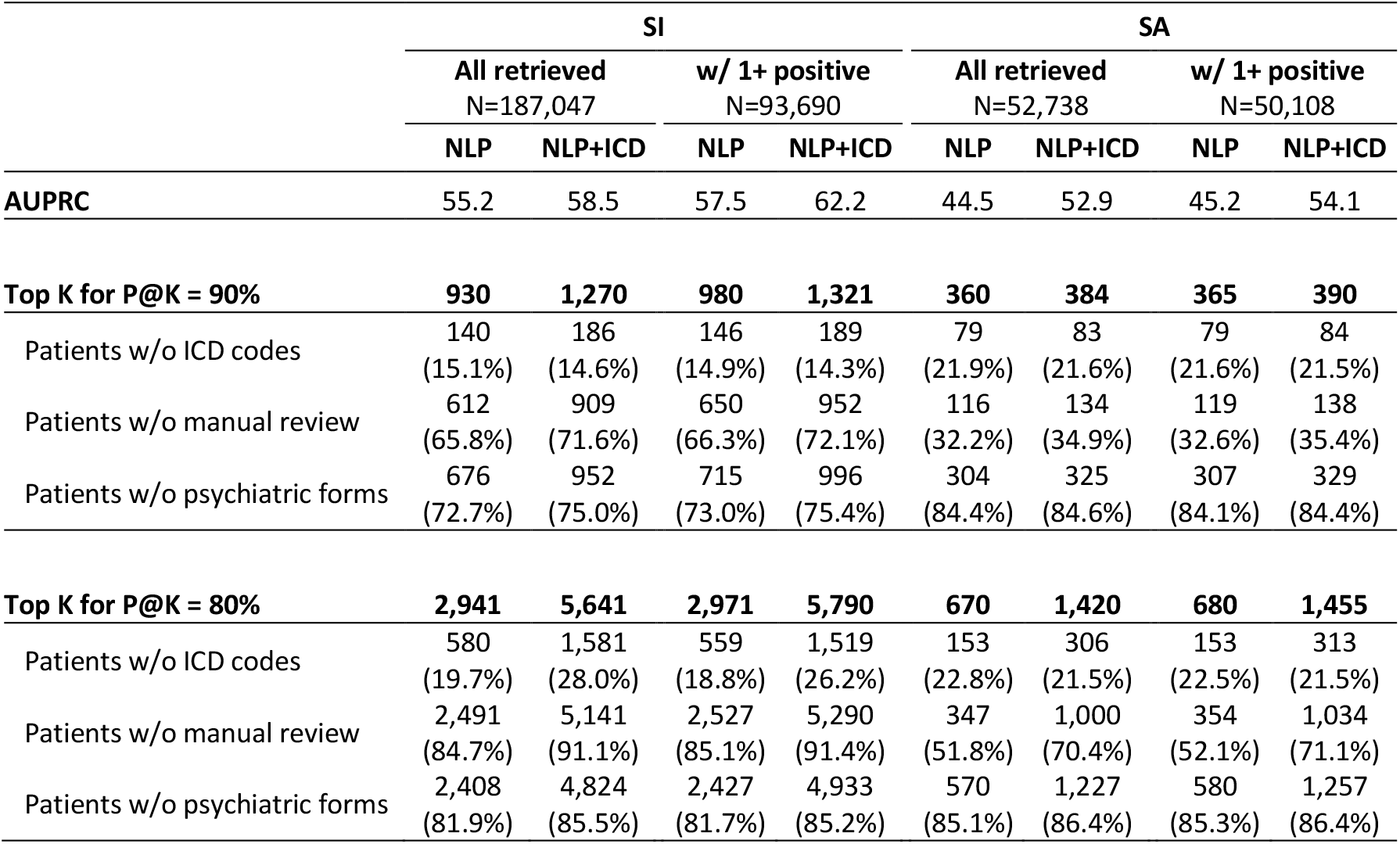
Evaluation of label assignment methods for suicidal ideation (SI) and suicide attempt (SA). The “All retrieved” columns represent results of the methods using the initial lists with all the retrieved patients for SI (N=187,047) and SA (N=52,738). The “w/ 1+ positive” columns correspond to methods using only patients with at least one positively asserted suicide mention in their notes. “NLP” and “NLP+ICD” columns are associated with methods using *p*_*rank*_ and *p*_*NLP+ICD*9_, respectively, for suicide label assignment.

**Figure 3.**
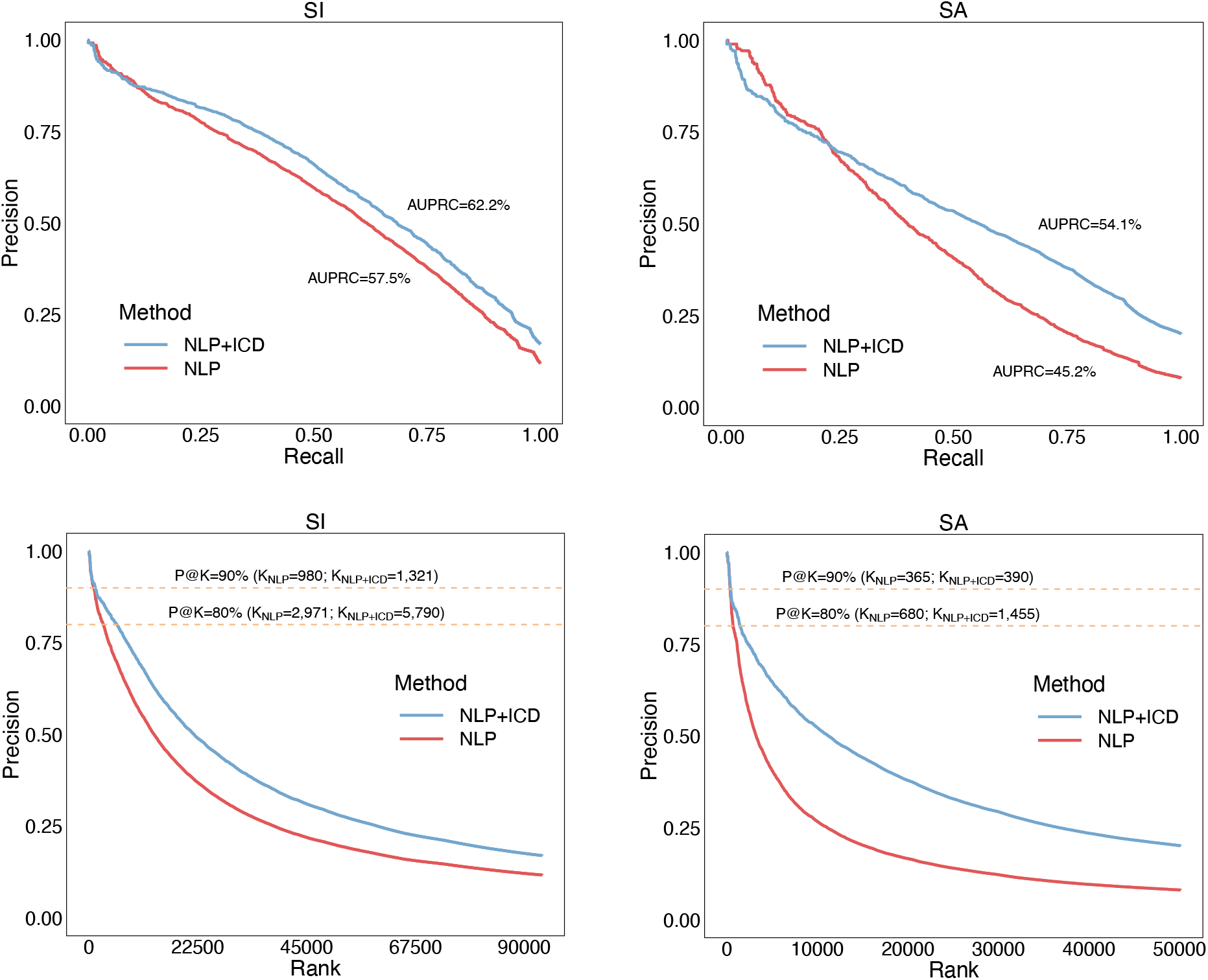
Evaluation comparing NLP and NLP+ICD label assignment methods for suicidal ideation (SI) and suicide attempt (SA). The patients used in this evaluation contain at least one positively asserted mention of suicide in their notes.

**Figure 4.**
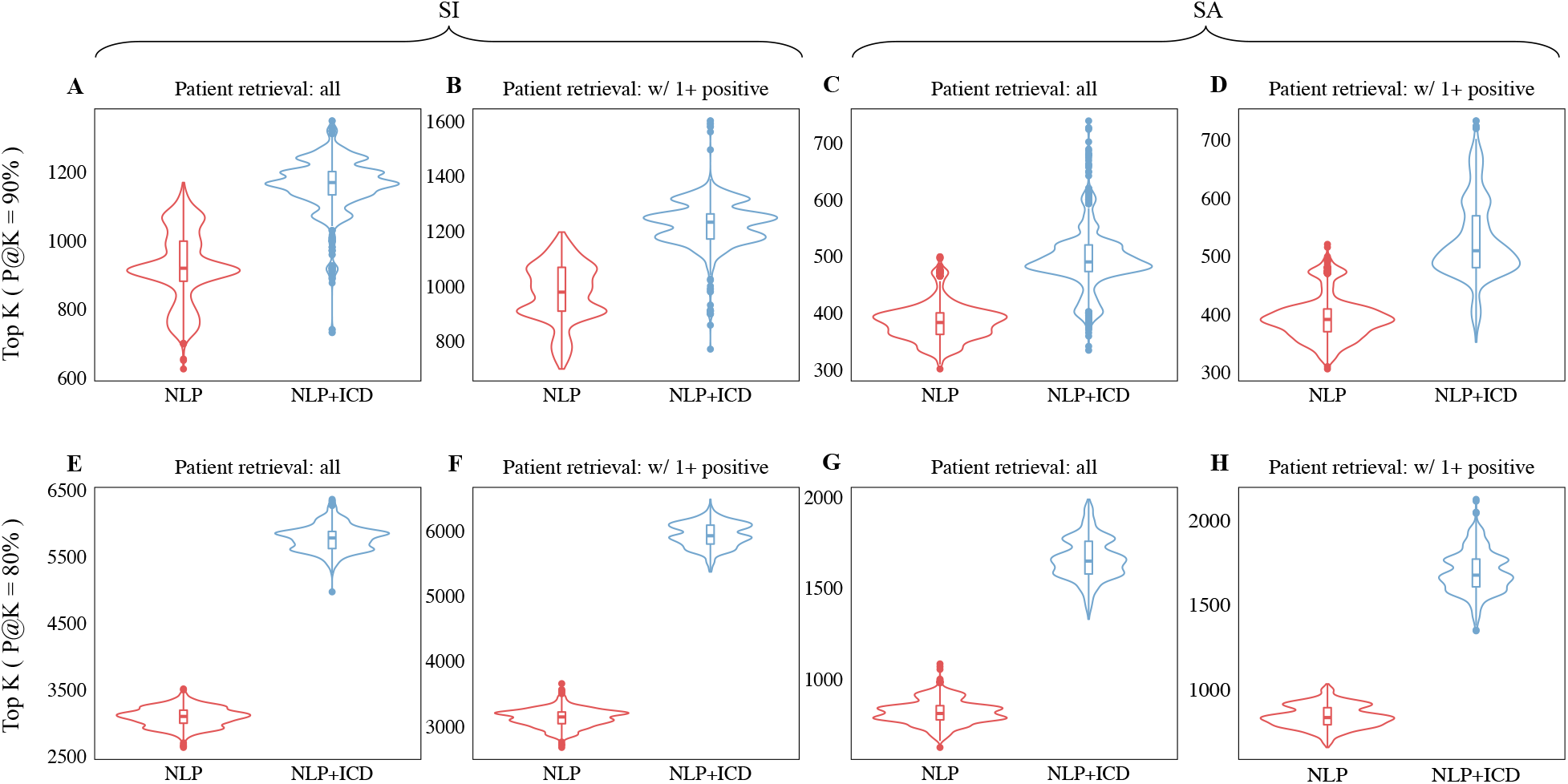
Comparative analysis for extracting the top K highest ranked suicidal ideation (SI) and suicide attempt (SA) patients using various configurations of the suicide label assignment method described in **Algorithm 1**. For each configuration, the label assignment method was run 1,000 times. The “Patient retrieval: all” experiments include all the patients retrieved by the NLP system while the “Patient retrieval: w/ 1+ positive” experiments use only patients with at least one positive suicide mention in their notes. The “NLP” and “NLP+ICD” experiments were associated with methods using the label assignment probabilities *p*_*rank*_ and *p*_*NLP+ICD*9_, respectively.

### High-precision extraction of suicidal ideation and suicide attempt cases

For case extraction, we selected the method that relied on both NLP ranks and ICD codes and included only the patients with at least one positive mention of suicide in their notes. As reported above, this method achieved the best AUPRC values and identified the highest number of cases in the top K highest ranked patients for P@K=90% and P@K=80%. Out of the suicidal ideation cases extracted with a precision of at least 90%, 72% (N=952) were automatically identified by the method (i.e., they were not manually reviewed) and 75% (N=996) did not have psychiatric forms for suicide assessment. Similarly, 71% (N=1,034) of the suicide attempt cases identified with a precision of at least 80% were automatically labeled (**Table 3**).

To extract cases with high precision from the entire EHR repository, in addition to the highest ranked patients identified by the NLP system, we included all other patients manually labeled as cases or with positive assertions for suicidal ideation and suicide attempt in their psychiatric forms. Based on the validation results achieved in this study, we also included all the patients with relevant ICD10CM codes. Specifically, the ICD10CM-based inclusion was performed for the extraction of suicidal ideation and suicide attempt cases with a precision/PPV of at least 96% and 85%, respectively. Conversely, from these cohorts, we excluded the patients that were manually labeled as non-cases or patients with negative assertions of the two suicide phenotypes in their psychiatric forms. For instance, to extract the suicidal ideation cases with a precision of at least 90%, we included the top 1,209 highest ranked patients in the corresponding list. The number of cases increased to 22,218 after including the cases derived from manual chart review and psychiatric forms as well as patients with relevant ICD10CM codes (**Table 4** and **Table S8**). Notably, most of these additional cases of suicidal ideation from **Table S8** were also included in the corresponding NLP list and were ranked below 1,209 (in this example, P@1,209=90%).

**Table 4.** High-precision extraction of suicidal ideation (SI) and suicide attempt (SA) cases extracted from the EHR.

| PPV criterion | SI |  | SA |  |
| --- | --- | --- | --- | --- |
|  | Resource | N (pooled) | Resource | N (pooled) |
| PPV ≥ 90% | NLP, top K, $P@K=90$ | 1,209 | NLP, top K, $P@K=90$ | 380 |
|  | + chart review (SI cases) | 1,831 | + chart review (SA cases) | 846 |
|  | + psychiatric forms (SI cases) | 6,670 | + psychiatric forms (SA cases) | **4,978** |
|  | + ICD10CM codes (PPV=96%) | **22,218** |  |  |
| PPV ≥ 80% | NLP, top K, $P@K=80$ | 5,342 | NLP, top K, $P@K=80$ | 1,384 |
|  | + chart review (SI cases) | 5,833 | + chart review (SA cases) | 1,681 |
|  | + psychiatric forms (SI cases) | 10,150 | + psychiatric forms (SA cases) | 5,701 |
|  | + ICD10CM codes (PPV=96%) | **23,848** | + ICD10CM codes (PPV=85%) | **18,843** |

The patients in this specific cohort of suicidal ideation cases were predominantly females (54.9%), White (77.5%), not Hispanic or Latino (89.8%), and with a mean age of 31.7 years (**Table 5**).

**Table 5.** Characteristics of suicidal ideation (SI) and suicide attempt (SA) cases identified in the EHR with a precision of at least 90%.

| Characteristic | SI |  | SA |  |
| --- | --- | --- | --- | --- |
|  | N | % | N | % |
| Total | 22,218 | 100 | 4,978 | 100 |
| Age, years* | 31.7 | 17.4 | 35.6 | 15.7 |
| Dead | 541 | 2.4 | 207 | 4.2 |
| **Sex** |  |  |  |  |
| Male | 10,026 | 45.1 | 2,165 | 43.5 |
| Female | 12,191 | 54.9 | 2,813 | 56.5 |
| Unknown | 1 | 0 | 0 | 0 |
| **Race** |  |  |  |  |
| White | 17,210 | 77.5 | 3,999 | 80.3 |
| Black | 3,254 | 14.6 | 743 | 14.9 |
| Asian | 271 | 1.2 | 37 | 0.7 |
| Native | 38 | 0.2 | 14 | 0.3 |
| Unknown | 1,445 | 6.5 | 185 | 3.7 |
| **Ethnicity** |  |  |  |  |
| Not Hispanic or Latino | 19,957 | 89.8 | 4,686 | 94.1 |
| Hispanic or Latino | 919 | 4.1 | 125 | 2.5 |
| Unknown | 1,342 | 6.0 | 167 | 3.4 |

\* Reported as mean and standard deviation

## DISCUSSION

This study developed, validated, and compared scalable NLP methodologies to ascertain prevalent suicidal ideation and suicide attempt from longitudinal EHRs. We relied on validated methods proven in other phenotypes in social determinants of health, and we compared NLP to diagnostic coding, the basis for other scalable ascertainment efforts. We emphasized positive predictive value/precision as a key metric of comparison – critically important in a domain characterized by rare events and the potential for stigma from false positives.^6,22^ Both ICD10CM-based and NLP-based ascertainment methods performed well, with NLP demonstrating consistently excellent PPV (> 95% for both outcomes). The increased granularity and specificity of ICD10CM (PPV=85%) likely contributed notably to improving on ICD9CM in prior chart validation on over 5,500 charts (PPV=58.63%).^4^ An ideal solution for ascertaining suicidal ideation and suicide attempt was provided by psychiatric forms when available in EHR. However, many centers like ours do not collect these data at scale^23^ or perform universal screening outside of emergency departments where it’s been shown to be feasible.^24-26^

Two key methodologies leveraging NLP and ICD codes were proposed for the extraction of suicidal ideation and suicide attempt cases. First, the selection of the top K highest ranked patients as cases was motivated by the NLP-based validation results. The motivation for the “top-K” approach was reinforced by the fact that 1) patients with ICD codes for self-injurious thoughts and behaviors (as compared to the patients without relevant ICD codes) achieved better performance for the NLP-based validation and 2) patients with relevant ICD codes are ranked towards the top of the retrieved patient lists. Second, the weakly supervised approach proposed for case label assignment enabled the selection of the most relevant cases from the entire EHR repository, all of whom being extracted with a precision above a prespecified lower bound.

Moreover, the weakly supervised approach allowed us to demonstrate that the results achieved by combining NLP ranks and ICD codes outperformed the results obtained by NLP ranks alone. When accounting for negation, we showed that excluding the patients with all their suicide mentions negated in their notes further increased the identification of suicidal ideation and suicide attempt cases. This improvement could be explained by a higher prevalence of patients with at least one positive mention of suicide (as compared to the prevalence of patients with all their suicide mentions negated) among 1) the manually validated cases, 2) the patients with positive assertions for suicidal ideation and suicide attempt in their psychiatric forms, and 3) the patients with relevant ICD codes. Interestingly, we also found that the proportion of patients with at least one positively asserted suicide mention in the suicide attempt list (95%) is substantially higher than the corresponding proportion in the suicidal ideation list (50%) suggesting that screening for these two phenotypes in notes is different.

The primary findings of this study support the applicability of NLP to additional phenotypes fraught with under-coding, under-reporting, and stigma.^27-29^ A well-powered chart validation supported the concurrent validity of this method not just for suicide attempt but also for a related phenotype, suicidal ideation. This NLP system can be applied to any unstructured clinical text common in EHRs and is feasible to apply at scale (∼200M notes here). This information retrieval approach would be portable to other health systems and has been used for the investigation of social determinants of health.^12,13^

This study was built on prior work by adding to our understanding of the validity of ICD codes to ascertain suicide phenotypes from EHRs. It added an NLP approach validated in other phenotypes to a novel application here for suicidal ideation and suicide attempt in order to inform both research and clinical operational work in this domain. Real-time clinical predictive studies are now underway and rely on structured data, e.g., diagnostic codes and problem lists, to track outcomes.^30^ Because of inherent delays in diagnostic coding, systems that might review clinical text as it is entered into EHRs would enable faster and more accurate ascertainment to inform learning health systems for suicide prevention.

Strengths of this study included reliance on a validated NLP approach with clinical expertise to guide query reformulation. We applied these methods to a large repositoty of real-world EHRs from a major academic medical center and conducted multi-expert, unbiased chart review for validation. We compared NLP to combinations of ICD codes and structured forms to ascertain suicidal ideation and suicide attempt with high precision.

Limitations of this study included its single study nature and potential for local coding and documentation practices that might not generalize to other healthcare systems. The chart validation focused on a “top-K” unbiased analysis which provided holistic understanding of the performance at the top ranked charts via NLP, but it provided less insight into potential uncertainty in precision for charts ranked with lower relevance scores. Our analyses emphasized *prevalent* suicidality – events throughout one’s lifetime – which are critical for precision medicine and phenotyping studies. However, clinically, *incident* detection systems are paramount. Longer term, we seek clinical systems that identify a new incident of, e.g., suicidal behavior, and differentiating a subsequent new event from a lifetime history or prior event remains a challenging ascertainment problem.

Future work should replicate this NLP approach both with and without suicide query reformulation in novel settings and on novel text corpora. Integrating systems incorporating structured and unstructured data streams requires subsequent implementation science and informatics efforts in this domain. Explainability and transparency of these algorithms will become more important as systems using them near clinical deployment.

## CONCLUSION

Scalable NLP based on information retrieval demonstrated high precision to identify suicidal ideation and suicide attempt across 200M clinical notes. A system leveraging both diagnostic coding and NLP might yield optimal ascertainment to inform phenotyping, clinical prediction, and monitoring applications in real-world healthcare systems. Future work should attempt to replicate these findings, should consider incident events in place of prevalent events, and should broach implementation needs for clinical and phenotyping research systems leveraging their potential to reach unprecedented quality and accuracy in ascertainment of suicidality.

## Data Availability

Data contain protected health information and are not publicly available. The summary statistics extracted from the EHR data used in this study are provided in the manuscript and supplementary material.

## Author contributions

C.A.B. and C.G.W. wrote the manuscript. C.A.B., D.M.R., and C.G.W. designed the research. C.A.B. implemented the NLP system. C.G.W. contributed to analytical tools. R.A, K.R. and C.G.W. performed the chart review. M.R., D.W., J.K., T.J.M., D.M.R., and C.G.W. provided critical suggestions and clinical insights into the analysis of suicide phenotypes. All authors contributed to the final manuscript.

