## Supplementary Material for "Improving ascertainment of suicidal ideation and suicide attempt with natural language processing"

### Supplemental Tables

**Table S1** ICD9CM codes for suicidal ideation.

**Table S2** ICD10CM codes for suicidal ideation.

**Table S3** ICD9CM codes for suicide attempt.

**Table S4** ICD10CM codes suicide attempt.

**Table S5** Top 50 keywords generated by word2vec as semantically similar to 'suicide' and 'suicidal'.

**Table S6** Query terms used to retrieve suicidal ideation and suicide attempt.

**Table S7** Comparative analysis for extracting the top K highest ranked suicidal ideation and suicide attempt patients using various configurations of the suicide label assignment method.

**Table S8** Suicidal ideation and suicide attempt cases extracted from the EHR.

**Table S1** ICD9CM codes for suicidal ideation.

| ICD9CM | Description |
|---|---|
| V62.84 | Suicidal ideation |

**Table S2** ICD10CM codes for suicidal ideation.

| ICD10CM | Description |
|---|---|
| R45.851 | Suicidal ideations |

**Table S3** ICD9CM codes for suicide attempt.

| ICD9CM | Description |
|---|---|
| E950* | Suicide and self-inflicted poisoning by solid or liquid substances |
| E951* | Suicide and self-inflicted poisoning by gases in domestic use |
| E952* | Suicide and self-inflicted poisoning by other gases and vapors |
| E953* | Suicide and self-inflicted injury by hanging, strangulation, and suffocation |
| E954 | Suicide and self-inflicted injury by submersion [drowning] |
| E955* | Suicide and self-inflicted injury by firearms, air guns, and explosives |
| E956 | Suicide and self-inflicted injury by cutting and piercing instrument |
| E957* | Suicide and self-inflicted injuries by jumping from high place |
| E958* | Suicide and self-inflicted injury by other and unspecified means |
| E959 | Late effects of self-inflicted injury |
| E980* | Poisoning by solid or liquid substances, undetermined whether accidentally or purposely inflicted |
| E981* | Poisoning by gases in domestic use, undetermined whether accidentally or purposely inflicted |
| E982* | Poisoning by other gases, undetermined whether accidentally or purposely inflicted |
| E983* | Hanging, strangulation, or suffocation, undetermined whether accidentally or purposely inflicted |
| E984 | Submersion (drowning), undetermined whether accidentally or purposely inflicted |
| E985* | Injury by firearms, air guns and explosives, undetermined whether accidentally or purposely inflicted |
| E986 | Injury by cutting and piercing instruments, undetermined whether accidentally or purposely inflicted |
| E987* | Falling from high place, undetermined whether accidentally or purposely inflicted |
| E988* | Injury by other and unspecified means, undetermined whether accidentally or purposely inflicted |
| E989 | Late effects of injury, undetermined whether accidentally or purposely inflicted |

**Table S4** ICD10CM codes suicide attempt.

| ICD10CM | Description |
|---|---|
| T14.91* | Suicide attempt |
| T36.0X2* | Poisoning by penicillins, intentional self-harm |
| T36.0X4* | Poisoning by penicillins, undetermined |
| T36.1X2* | Poisoning by cephalosporins and other beta-lactam antibiotics, intentional self-harm |
| T36.1X4* | Poisoning by cephalosporins and other beta-lactam antibiotics, undetermined |
| T36.2X2* | Poisoning by chloramphenicol group, intentional self-harm |
| T36.2X4* | Poisoning by chloramphenicol group, undetermined |
| T36.3X2* | Poisoning by macrolides, intentional self-harm |
| T36.3X4* | Poisoning by macrolides, undetermined |
| T36.4X2* | Poisoning by tetracyclines, intentional self-harm |
| T36.4X4* | Poisoning by tetracyclines, undetermined |
| T36.5X2* | Poisoning by aminoglycosides, intentional self-harm |
| T36.5X4* | Poisoning by aminoglycosides, undetermined |
| T36.6X2* | Poisoning by rifampicins, intentional self-harm |
| T36.6X4* | Poisoning by rifampicins, undetermined |

| | |
|---|---|
| T36.7X2* | Poisoning by antifungal antibiotics, systemically used, intentional self-harm |
| T36.7X4* | Poisoning by antifungal antibiotics, systemically used, undetermined |
| T36.8X2* | Poisoning by other systemic antibiotics, intentional self-harm |
| T36.8X4* | Poisoning by other systemic antibiotics, undetermined |
| T36.92* | Poisoning by unspecified systemic antibiotic, intentional self-harm |
| T36.94* | Poisoning by unspecified systemic antibiotic, undetermined |
| T37.0X2* | Poisoning by sulfonamides, intentional self-harm |
| T37.0X4* | Poisoning by sulfonamides, undetermined |
| T37.1X2* | Poisoning by antimycobacterial drugs, intentional self-harm |
| T37.1X4* | Poisoning by antimycobacterial drugs, undetermined |
| T37.2X2* | Poisoning by antimalarials and drugs acting on other blood protozoa, intentional self-harm |
| T37.2X4* | Poisoning by antimalarials and drugs acting on other blood protozoa, undetermined |
| T37.3X2* | Poisoning by other antiprotozoal drugs, intentional self-harm |
| T37.3X4* | Poisoning by other antiprotozoal drugs, undetermined |
| T37.4X2* | Poisoning by anthelminthics, intentional self-harm |
| T37.4X4* | Poisoning by anthelminthics, undetermined |
| T37.5X2* | Poisoning by antiviral drugs, intentional self-harm |
| T37.5X4* | Poisoning by antiviral drugs, undetermined |
| T37.8X2* | Poisoning by other specified systemic anti-infectives and antiparasitics, intentional self-harm |
| T37.8X4* | Poisoning by other specified systemic anti-infectives and antiparasitics, undetermined |
| T37.92* | Poisoning by unspecified systemic anti-infective and antiparasitics, intentional self-harm |
| T37.94* | Poisoning by unspecified systemic anti-infective and antiparasitics, undetermined |
| T38.0X2* | Poisoning by glucocorticoids and synthetic analogues, intentional self-harm |
| T38.0X4* | Poisoning by glucocorticoids and synthetic analogues, undetermined |
| T38.1X2* | Poisoning by thyroid hormones and substitutes, intentional self-harm |
| T38.1X4* | Poisoning by thyroid hormones and substitutes, undetermined |
| T38.2X2* | Poisoning by antithyroid drugs, intentional self-harm |
| T38.2X4* | Poisoning by antithyroid drugs, undetermined |
| T38.3X2* | Poisoning by insulin and oral hypoglycemic [antidiabetic] drugs, intentional self-harm |
| T38.3X4* | Poisoning by insulin and oral hypoglycemic [antidiabetic] drugs, undetermined |
| T38.4X2* | Poisoning by oral contraceptives, intentional self-harm |
| T38.4X4* | Poisoning by oral contraceptives, undetermined |
| T38.5X2* | Poisoning by other estrogens and progestogens, intentional self-harm |
| T38.5X4* | Poisoning by other estrogens and progestogens, undetermined |
| T38.6X2* | Poisoning by antigonadotrophins, antiestrogens, antiandrogens, not elsewhere classified, intentional self-harm |
| T38.6X4* | Poisoning by antigonadotrophins, antiestrogens, antiandrogens, not elsewhere classified, undetermined |
| T38.7X2* | Poisoning by androgens and anabolic congeners, intentional self-harm |
| T38.7X4* | Poisoning by androgens and anabolic congeners, undetermined |
| T38.802* | Poisoning by unspecified hormones and synthetic substitutes, intentional self-harm |
| T38.804* | Poisoning by unspecified hormones and synthetic substitutes, undetermined |
| T38.812* | Poisoning by anterior pituitary [adenohypophyseal] hormones, intentional self-harm |
| T38.814* | Poisoning by anterior pituitary [adenohypophyseal] hormones, undetermined |
| T38.892* | Poisoning by other hormones and synthetic substitutes, intentional self-harm |
| T38.894* | Poisoning by other hormones and synthetic substitutes, undetermined |
| T38.902* | Poisoning by unspecified hormone antagonists, intentional self-harm |
| T38.904* | Poisoning by unspecified hormone antagonists, undetermined |
| T38.992* | Poisoning by other hormone antagonists, intentional self-harm |
| T38.994* | Poisoning by other hormone antagonists, undetermined |
| T39.012* | Poisoning by aspirin, intentional self-harm |
| T39.014* | Poisoning by aspirin, undetermined |
| T39.092* | Poisoning by salicylates, intentional self-harm |

| | |
|---|---|
| T39.094* | Poisoning by salicylates, undetermined |
| T39.1X2* | Poisoning by 4-Aminophenol derivatives, intentional self-harm |
| T39.1X4* | Poisoning by 4-Aminophenol derivatives, undetermined |
| T39.2X2* | Poisoning by pyrazolone derivatives, intentional self-harm |
| T39.2X4* | Poisoning by pyrazolone derivatives, undetermined |
| T39.312* | Poisoning by propionic acid derivatives, intentional self-harm |
| T39.314* | Poisoning by propionic acid derivatives, undetermined |
| T39.392* | Poisoning by other nonsteroidal anti-inflammatory drugs [NSAID], intentional self-harm |
| T39.394* | Poisoning by other nonsteroidal anti-inflammatory drugs [NSAID], undetermined |
| T39.4X2* | Poisoning by antirheumatics, not elsewhere classified, intentional self-harm |
| T39.4X4* | Poisoning by antirheumatics, not elsewhere classified, undetermined |
| T39.8X2* | Poisoning by other nonopioid analgesics and antipyretics, not elsewhere classified, intentional self-harm |
| T39.8X4* | Poisoning by other nonopioid analgesics and antipyretics, not elsewhere classified, undetermined |
| T39.92* | Poisoning by unspecified nonopioid analgesic, antipyretic and antirheumatic, intentional self-harm |
| T39.94* | Poisoning by unspecified nonopioid analgesic, antipyretic and antirheumatic, undetermined |
| T40.0X2* | Poisoning by opium, intentional self-harm |
| T40.0X4* | Poisoning by opium, undetermined |
| T40.1X2* | Poisoning by heroin, intentional self-harm |
| T40.1X4* | Poisoning by heroin, undetermined |
| T40.2X2* | Poisoning by other opioids, intentional self-harm |
| T40.2X4* | Poisoning by other opioids, undetermined |
| T40.3X2* | Poisoning by methadone, intentional self-harm |
| T40.3X4* | Poisoning by methadone, undetermined |
| T40.4X2* | Poisoning by other synthetic narcotics, intentional self-harm |
| T40.4X4* | Poisoning by other synthetic narcotics, undetermined |
| T40.5X2* | Poisoning by cocaine, intentional self-harm |
| T40.5X4* | Poisoning by cocaine, undetermined |
| T40.602* | Poisoning by unspecified narcotics, intentional self-harm |
| T40.604* | Poisoning by unspecified narcotics, undetermined |
| T40.692* | Poisoning by other narcotics, intentional self-harm |
| T40.694* | Poisoning by other narcotics, undetermined |
| T40.7X2* | Poisoning by cannabis (derivatives), intentional self-harm |
| T40.7X4* | Poisoning by cannabis (derivatives), undetermined |
| T40.8X2* | Poisoning by lysergide [LSD], intentional self-harm |
| T40.8X4* | Poisoning by lysergide [LSD], undetermined |
| T40.902* | Poisoning by unspecified psychodysleptics [hallucinogens], intentional self-harm |
| T40.904* | Poisoning by unspecified psychodysleptics [hallucinogens], undetermined |
| T40.992* | Poisoning by other psychodysleptics [hallucinogens], intentional self-harm |
| T40.994* | Poisoning by other psychodysleptics [hallucinogens], undetermined |
| T41.0X2* | Poisoning by inhaled anesthetics, intentional self-harm |
| T41.0X4* | Poisoning by inhaled anesthetics, undetermined |
| T41.1X2* | Poisoning by intravenous anesthetics, intentional self-harm |
| T41.1X4* | Poisoning by intravenous anesthetics, undetermined |
| T41.202* | Poisoning by unspecified general anesthetics, intentional self-harm |
| T41.204* | Poisoning by unspecified general anesthetics, undetermined |
| T41.292* | Poisoning by other general anesthetics, intentional self-harm |
| T41.294* | Poisoning by other general anesthetics, undetermined |
| T41.3X2* | Poisoning by local anesthetics, intentional self-harm |
| T41.3X4* | Poisoning by local anesthetics, undetermined |
| T41.42* | Poisoning by unspecified anesthetic, intentional self-harm |
| T41.44* | Poisoning by unspecified anesthetic, undetermined |
| T41.5X2* | Poisoning by therapeutic gases, intentional self-harm |

| | |
|---|---|
| T41.5X4* | Poisoning by therapeutic gases, undetermined |
| T42.0X2* | Poisoning by hydantoin derivatives, intentional self-harm |
| T42.0X4* | Poisoning by hydantoin derivatives, undetermined |
| T42.1X2* | Poisoning by iminostilbenes, intentional self-harm |
| T42.1X4* | Poisoning by iminostilbenes, undetermined |
| T42.2X2* | Poisoning by succinimides and oxazolidinediones, intentional self-harm |
| T42.2X4* | Poisoning by succinimides and oxazolidinediones, undetermined |
| T42.3X2* | Poisoning by barbiturates, intentional self-harm |
| T42.3X4* | Poisoning by barbiturates, undetermined |
| T42.4X2* | Poisoning by benzodiazepines, intentional self-harm |
| T42.4X4* | Poisoning by benzodiazepines, undetermined |
| T42.5X2* | Poisoning by mixed antiepileptics, intentional self-harm |
| T42.5X4* | Poisoning by mixed antiepileptics, undetermined |
| T42.6X2* | Poisoning by other antiepileptic and sedative-hypnotic drugs, intentional self-harm |
| T42.6X4* | Poisoning by other antiepileptic and sedative-hypnotic drugs, undetermined |
| T42.72* | Poisoning by unspecified antiepileptic and sedative-hypnotic drugs, intentional self-harm |
| T42.74* | Poisoning by unspecified antiepileptic and sedative-hypnotic drugs, undetermined |
| T42.8X2* | Poisoning by antiparkinsonism drugs and other central muscle-tone depressants, intentional self-harm |
| T42.8X4* | Poisoning by antiparkinsonism drugs and other central muscle-tone depressants, undetermined |
| T43.012* | Poisoning by tricyclic antidepressants, intentional self-harm |
| T43.014* | Poisoning by tricyclic antidepressants, undetermined |
| T43.022* | Poisoning by tetracyclic antidepressants, intentional self-harm |
| T43.024* | Poisoning by tetracyclic antidepressants, undetermined |
| T43.0X2* | Poisoning by tricyclic and tetracyclic antidepressants, intentional self-harm |
| T43.0X4* | Poisoning by tricyclic and tetracyclic antidepressants, undetermined |
| T43.1X2* | Poisoning by monoamine-oxidase-inhibitor antidepressants, intentional self-harm |
| T43.1X4* | Poisoning by monoamine-oxidase-inhibitor antidepressants, undetermined |
| T43.202* | Poisoning by unspecified antidepressants, intentional self-harm |
| T43.204* | Poisoning by unspecified antidepressants, undetermined |
| T43.212* | Poisoning by selective serotonin and norepinephrine reuptake inhibitors, intentional self-harm |
| T43.214* | Poisoning by selective serotonin and norepinephrine reuptake inhibitors, undetermined |
| T43.222* | Poisoning by selective serotonin reuptake inhibitors, intentional self-harm |
| T43.224* | Poisoning by selective serotonin reuptake inhibitors, undetermined |
| T43.292* | Poisoning by other antidepressants, intentional self-harm |
| T43.294* | Poisoning by other antidepressants, undetermined |
| T43.3X2* | Poisoning by phenothiazine antipsychotics and neuroleptics, intentional self-harm |
| T43.3X4* | Poisoning by phenothiazine antipsychotics and neuroleptics, undetermined |
| T43.4X2* | Poisoning by butyrophenone and thiothixene neuroleptics, intentional self-harm |
| T43.4X4* | Poisoning by butyrophenone and thiothixene neuroleptics, undetermined |
| T43.502* | Poisoning by unspecified antipsychotics and neuroleptics, intentional self-harm |
| T43.504* | Poisoning by unspecified antipsychotics and neuroleptics, undetermined |
| T43.592* | Poisoning by other antipsychotics and neuroleptics, intentional self-harm |
| T43.594* | Poisoning by other antipsychotics and neuroleptics, undetermined |
| T43.602* | Poisoning by unspecified psychostimulants, intentional self-harm |
| T43.604* | Poisoning by unspecified psychostimulants, undetermined |
| T43.612* | Poisoning by caffeine, intentional self-harm |
| T43.614* | Poisoning by caffeine, undetermined |
| T43.622* | Poisoning by amphetamines, intentional self-harm |
| T43.624* | Poisoning by amphetamines, undetermined |
| T43.632* | Poisoning by methylphenidate, intentional self-harm |
| T43.634* | Poisoning by methylphenidate, undetermined |
| T43.642* | Poisoning by ecstasy, intentional self-harm |

| | |
|---|---|
| T43.644* | Poisoning by ecstasy, undetermined |
| T43.692* | Poisoning by other psychostimulants, intentional self-harm |
| T43.694* | Poisoning by other psychostimulants, undetermined |
| T43.6X2* | Poisoning by psychostimulants with abuse potential, intentional self-harm |
| T43.6X4* | Poisoning by psychostimulants with abuse potential, undetermined |
| T43.8X2* | Poisoning by other psychotropic drugs, intentional self-harm |
| T43.8X4* | Poisoning by other psychotropic drugs, undetermined |
| T43.92* | Poisoning by unspecified psychotropic drug, intentional self-harm |
| T43.94* | Poisoning by unspecified psychotropic drug, undetermined |
| T44.0X2* | Poisoning by anticholinesterase agents, intentional self-harm |
| T44.0X4* | Poisoning by anticholinesterase agents, undetermined |
| T44.1X2* | Poisoning by other parasympathomimetics [cholinergics], intentional self-harm |
| T44.1X4* | Poisoning by other parasympathomimetics [cholinergics], undetermined |
| T44.2X2* | Poisoning by ganglionic blocking drugs, intentional self-harm |
| T44.2X4* | Poisoning by ganglionic blocking drugs, undetermined |
| T44.3X2* | Poisoning by other parasympatholytics [anticholinergics and antimuscarinics] and spasmolytics, intentional self-harm |
| T44.3X4* | Poisoning by other parasympatholytics [anticholinergics and antimuscarinics] and spasmolytics, undetermined |
| T44.4X2* | Poisoning by predominantly alpha-adrenoreceptor agonists, intentional self-harm |
| T44.4X4* | Poisoning by predominantly alpha-adrenoreceptor agonists, undetermined |
| T44.5X2* | Poisoning by predominantly beta-adrenoreceptor agonists, intentional self-harm |
| T44.5X4* | Poisoning by predominantly beta-adrenoreceptor agonists, undetermined |
| T44.6X2* | Poisoning by alpha-adrenoreceptor antagonists, intentional self-harm |
| T44.6X4* | Poisoning by alpha-adrenoreceptor antagonists, undetermined |
| T44.7X2* | Poisoning by beta-adrenoreceptor antagonists, intentional self-harm |
| T44.7X4* | Poisoning by beta-adrenoreceptor antagonists, undetermined |
| T44.8X2* | Poisoning by centrally-acting and adrenergic-neuron-blocking agents, intentional self-harm |
| T44.8X4* | Poisoning by centrally-acting and adrenergic-neuron-blocking agents, undetermined |
| T44.902* | Poisoning by unspecified drugs primarily affecting the autonomic nervous system, intentional self-harm |
| T44.904* | Poisoning by unspecified drugs primarily affecting the autonomic nervous system, undetermined |
| T44.992* | Poisoning by other drug primarily affecting the autonomic nervous system, intentional self-harm |
| T44.994* | Poisoning by other drug primarily affecting the autonomic nervous system, undetermined |
| T45.0X2* | Poisoning by antiallergic and antiemetic drugs, intentional self-harm |
| T45.0X4* | Poisoning by antiallergic and antiemetic drugs, undetermined |
| T45.1X2* | Poisoning by antineoplastic and immunosuppressive drugs, intentional self-harm |
| T45.1X4* | Poisoning by antineoplastic and immunosuppressive drugs, undetermined |
| T45.2X2* | Poisoning by vitamins, intentional self-harm |
| T45.2X4* | Poisoning by vitamins, undetermined |
| T45.3X2* | Poisoning by enzymes, intentional self-harm |
| T45.3X4* | Poisoning by enzymes, undetermined |
| T45.4X2* | Poisoning by iron and its compounds, intentional self-harm |
| T45.4X4* | Poisoning by iron and its compounds, undetermined |
| T45.512* | Poisoning by anticoagulants, intentional self-harm |
| T45.514* | Poisoning by anticoagulants, undetermined |
| T45.522* | Poisoning by antithrombotic drugs, intentional self-harm |
| T45.524* | Poisoning by antithrombotic drugs, undetermined |
| T45.602* | Poisoning by unspecified fibrinolysis-affecting drugs, intentional self-harm |
| T45.604* | Poisoning by unspecified fibrinolysis-affecting drugs, undetermined |
| T45.612* | Poisoning by thrombolytic drug, intentional self-harm |
| T45.614* | Poisoning by thrombolytic drug, undetermined |
| T45.622* | Poisoning by hemostatic drug, intentional self-harm |

| | |
|---|---|
| T45.624* | Poisoning by hemostatic drug, undetermined |
| T45.692* | Poisoning by other fibrinolysis-affecting drugs, intentional self-harm |
| T45.694* | Poisoning by other fibrinolysis-affecting drugs, undetermined |
| T45.7X2* | Poisoning by anticoagulant antagonists, vitamin K and other coagulants, intentional self-harm |
| T45.7X4* | Poisoning by anticoagulant antagonists, vitamin K and other coagulants, undetermined |
| T45.8X2* | Poisoning by other primarily systemic and hematological agents, intentional self-harm |
| T45.8X4* | Poisoning by other primarily systemic and hematological agents, undetermined |
| T45.92* | Poisoning by unspecified primarily systemic and hematological agent, intentional self-harm |
| T45.94* | Poisoning by unspecified primarily systemic and hematological agent, undetermined |
| T46.0X2* | Poisoning by cardiac-stimulant glycosides and drugs of similar action, intentional self-harm |
| T46.0X4* | Poisoning by cardiac-stimulant glycosides and drugs of similar action, undetermined |
| T46.1X2* | Poisoning by calcium-channel blockers, intentional self-harm |
| T46.1X4* | Poisoning by calcium-channel blockers, undetermined |
| T46.2X2* | Poisoning by other antidysrhythmic drugs, intentional self-harm |
| T46.2X4* | Poisoning by other antidysrhythmic drugs, undetermined |
| T46.3X2* | Poisoning by coronary vasodilators, intentional self-harm |
| T46.3X4* | Poisoning by coronary vasodilators, undetermined |
| T46.4X2* | Poisoning by angiotensin-converting-enzyme inhibitors, intentional self-harm |
| T46.4X4* | Poisoning by angiotensin-converting-enzyme inhibitors, undetermined |
| T46.5X2* | Poisoning by other antihypertensive drugs, intentional self-harm |
| T46.5X4* | Poisoning by other antihypertensive drugs, undetermined |
| T46.6X2* | Poisoning by antihyperlipidemic and antiarteriosclerotic drugs, intentional self-harm |
| T46.6X4* | Poisoning by antihyperlipidemic and antiarteriosclerotic drugs, undetermined |
| T46.7X2* | Poisoning by peripheral vasodilators, intentional self-harm |
| T46.7X4* | Poisoning by peripheral vasodilators, undetermined |
| T46.8X2* | Poisoning by antivaricose drugs, including sclerosing agents, intentional self-harm |
| T46.8X4* | Poisoning by antivaricose drugs, including sclerosing agents, undetermined |
| T46.902* | Poisoning by unspecified agents primarily affecting the cardiovascular system, intentional self-harm |
| T46.904* | Poisoning by unspecified agents primarily affecting the cardiovascular system, undetermined |
| T46.992* | Poisoning by other agents primarily affecting the cardiovascular system, intentional self-harm |
| T46.994* | Poisoning by other agents primarily affecting the cardiovascular system, undetermined |
| T47.0X2* | Poisoning by histamine H2-receptor blockers, intentional self-harm |
| T47.0X4* | Poisoning by histamine H2-receptor blockers, undetermined |
| T47.1X2* | Poisoning by other antacids and anti-gastric-secretion drugs, intentional self-harm |
| T47.1X4* | Poisoning by other antacids and anti-gastric-secretion drugs, undetermined |
| T47.2X2* | Poisoning by stimulant laxatives, intentional self-harm |
| T47.2X4* | Poisoning by stimulant laxatives, undetermined |
| T47.3X2* | Poisoning by saline and osmotic laxatives, intentional self-harm |
| T47.3X4* | Poisoning by saline and osmotic laxatives, undetermined |
| T47.4X2* | Poisoning by other laxatives, intentional self-harm |
| T47.4X4* | Poisoning by other laxatives, undetermined |
| T47.5X2* | Poisoning by digestants, intentional self-harm |
| T47.5X4* | Poisoning by digestants, undetermined |
| T47.6X2* | Poisoning by antidiarrheal drugs, intentional self-harm |
| T47.6X4* | Poisoning by antidiarrheal drugs, undetermined |
| T47.7X2* | Poisoning by emetics, intentional self-harm |
| T47.7X4* | Poisoning by emetics, undetermined |
| T47.8X2* | Poisoning by other agents primarily affecting gastrointestinal system, intentional self-harm |
| T47.8X4* | Poisoning by other agents primarily affecting gastrointestinal system, undetermined |
| T47.92* | Poisoning by unspecified agents primarily affecting the gastrointestinal system, intentional self-harm |
| T47.94* | Poisoning by unspecified agents primarily affecting the gastrointestinal system, undetermined |
| T48.0X2* | Poisoning by oxytocic drugs, intentional self-harm |

| | |
|---|---|
| T48.0X4* | Poisoning by oxytocic drugs, undetermined |
| T48.1X2* | Poisoning by skeletal muscle relaxants [neuromuscular blocking agents], intentional self-harm |
| T48.1X4* | Poisoning by skeletal muscle relaxants [neuromuscular blocking agents], undetermined |
| T48.202* | Poisoning by unspecified drugs acting on muscles, intentional self-harm |
| T48.204* | Poisoning by unspecified drugs acting on muscles, undetermined |
| T48.292* | Poisoning by other drugs acting on muscles, intentional self-harm |
| T48.294* | Poisoning by other drugs acting on muscles, undetermined |
| T48.3X2* | Poisoning by antitussives, intentional self-harm |
| T48.3X4* | Poisoning by antitussives, undetermined |
| T48.4X2* | Poisoning by expectorants, intentional self-harm |
| T48.4X4* | Poisoning by expectorants, undetermined |
| T48.5X2* | Poisoning by other anti-common-cold drugs, intentional self-harm |
| T48.5X4* | Poisoning by other anti-common-cold drugs, undetermined |
| T48.6X2* | Poisoning by antiasthmatics, intentional self-harm |
| T48.6X4* | Poisoning by antiasthmatics, undetermined |
| T48.902* | Poisoning by unspecified agents primarily acting on the respiratory system, intentional self-harm |
| T48.904* | Poisoning by unspecified agents primarily acting on the respiratory system, undetermined |
| T48.992* | Poisoning by other agents primarily acting on the respiratory system, intentional self-harm |
| T48.994* | Poisoning by other agents primarily acting on the respiratory system, undetermined |
| T49.0X2* | Poisoning by local antifungal, anti-infective and anti-inflammatory drugs, intentional self-harm |
| T49.0X4* | Poisoning by local antifungal, anti-infective and anti-inflammatory drugs, undetermined |
| T49.1X2* | Poisoning by antipruritics, intentional self-harm |
| T49.1X4* | Poisoning by antipruritics, undetermined |
| T49.2X2* | Poisoning by local astringents and local detergents, intentional self-harm |
| T49.2X4* | Poisoning by local astringents and local detergents, undetermined |
| T49.3X2* | Poisoning by emollients, demulcents and protectants, intentional self-harm |
| T49.3X4* | Poisoning by emollients, demulcents and protectants, undetermined |
| T49.4X2* | Poisoning by keratolytics, keratoplastics, and other hair treatment drugs and preparations, intentional self-harm |
| T49.4X4* | Poisoning by keratolytics, keratoplastics, and other hair treatment drugs and preparations, undetermined |
| T49.5X2* | Poisoning by ophthalmological drugs and preparations, intentional self-harm |
| T49.5X4* | Poisoning by ophthalmological drugs and preparations, undetermined |
| T49.6X2* | Poisoning by otorhinolaryngological drugs and preparations, intentional self-harm |
| T49.6X4* | Poisoning by otorhinolaryngological drugs and preparations, undetermined |
| T49.7X2* | Poisoning by dental drugs, topically applied, intentional self-harm |
| T49.7X4* | Poisoning by dental drugs, topically applied, undetermined |
| T49.8X2* | Poisoning by other topical agents, intentional self-harm |
| T49.8X4* | Poisoning by other topical agents, undetermined |
| T49.92* | Poisoning by unspecified topical agent, intentional self-harm |
| T49.94* | Poisoning by unspecified topical agent, undetermined |
| T50.0X2* | Poisoning by mineralocorticoids and their antagonists, intentional self-harm |
| T50.0X4* | Poisoning by mineralocorticoids and their antagonists, undetermined |
| T50.1X2* | Poisoning by loop [high-ceiling] diuretics, intentional self-harm |
| T50.1X4* | Poisoning by loop [high-ceiling] diuretics, undetermined |
| T50.2X2* | Poisoning by carbonic-anhydrase inhibitors, benzothiadiazides and other diuretics, intentional self-harm |
| T50.2X4* | Poisoning by carbonic-anhydrase inhibitors, benzothiadiazides and other diuretics, undetermined |
| T50.3X2* | Poisoning by electrolytic, caloric and water-balance agents, intentional self-harm |
| T50.3X4* | Poisoning by electrolytic, caloric and water-balance agents, undetermined |
| T50.4X2* | Poisoning by drugs affecting uric acid metabolism, intentional self-harm |
| T50.4X4* | Poisoning by drugs affecting uric acid metabolism, undetermined |
| T50.5X2* | Poisoning by appetite depressants, intentional self-harm |

| | |
|---|---|
| T50.5X4* | Poisoning by appetite depressants, undetermined |
| T50.6X2* | Poisoning by antidotes and chelating agents, intentional self-harm |
| T50.6X4* | Poisoning by antidotes and chelating agents, undetermined |
| T50.7X2* | Poisoning by analeptics and opioid receptor antagonists, intentional self-harm |
| T50.7X4* | Poisoning by analeptics and opioid receptor antagonists, undetermined |
| T50.8X2* | Poisoning by diagnostic agents, intentional self-harm |
| T50.8X4* | Poisoning by diagnostic agents, undetermined |
| T50.902* | Poisoning by unspecified drugs, medicaments and biological substances, intentional self-harm |
| T50.904* | Poisoning by unspecified drugs, medicaments and biological substances, undetermined |
| T50.992* | Poisoning by other drugs, medicaments and biological substances, intentional self-harm |
| T50.994* | Poisoning by other drugs, medicaments and biological substances, undetermined |
| T50.A12* | Poisoning by pertussis vaccine, including combinations with a pertussis component, intentional self-harm |
| T50.A14* | Poisoning by pertussis vaccine, including combinations with a pertussis component, undetermined |
| T50.A22* | Poisoning by mixed bacterial vaccines without a pertussis component, intentional self-harm |
| T50.A24* | Poisoning by mixed bacterial vaccines without a pertussis component, undetermined |
| T50.A92* | Poisoning by other bacterial vaccines, intentional self-harm |
| T50.A94* | Poisoning by other bacterial vaccines, undetermined |
| T50.B12* | Poisoning by smallpox vaccines, intentional self-harm |
| T50.B14* | Poisoning by smallpox vaccines, undetermined |
| T50.B92* | Poisoning by other viral vaccines, intentional self-harm |
| T50.B94* | Poisoning by other viral vaccines, undetermined |
| T50.Z12* | Poisoning by immunoglobulin, intentional self-harm |
| T50.Z14* | Poisoning by immunoglobulin, undetermined |
| T50.Z92* | Poisoning by other vaccines and biological substances, intentional self-harm |
| T50.Z94* | Poisoning by other vaccines and biological substances, undetermined |
| T51.0X2* | Toxic effect of ethanol, intentional self-harm |
| T51.0X4* | Toxic effect of ethanol, undetermined |
| T51.1X2* | Toxic effect of methanol, intentional self-harm |
| T51.1X4* | Toxic effect of methanol, undetermined |
| T51.2X2* | Toxic effect of 2-Propanol, intentional self-harm |
| T51.2X4* | Toxic effect of 2-Propanol, undetermined |
| T51.3X2* | Toxic effect of fusel oil, intentional self-harm |
| T51.3X4* | Toxic effect of fusel oil, undetermined |
| T51.8X2* | Toxic effect of other alcohols, intentional self-harm |
| T51.8X4* | Toxic effect of other alcohols, undetermined |
| T51.92* | Toxic effect of unspecified alcohol, intentional self-harm |
| T51.94* | Toxic effect of unspecified alcohol, undetermined |
| T52.0X2* | Toxic effect of petroleum products, intentional self-harm |
| T52.0X4* | Toxic effect of petroleum products, undetermined |
| T52.1X2* | Toxic effect of benzene, intentional self-harm |
| T52.1X4* | Toxic effect of benzene, undetermined |
| T52.2X2* | Toxic effect of homologues of benzene, intentional self-harm |
| T52.2X4* | Toxic effect of homologues of benzene, undetermined |
| T52.3X2* | Toxic effect of glycols, intentional self-harm |
| T52.3X4* | Toxic effect of glycols, undetermined |
| T52.4X2* | Toxic effect of ketones, intentional self-harm |
| T52.4X4* | Toxic effect of ketones, undetermined |
| T52.8X2* | Toxic effect of other organic solvents, intentional self-harm |
| T52.8X4* | Toxic effect of other organic solvents, undetermined |
| T52.92* | Toxic effect of unspecified organic solvent, intentional self-harm |
| T52.94* | Toxic effect of unspecified organic solvent, undetermined |
| T53.0X2* | Toxic effect of carbon tetrachloride, intentional self-harm |

| | |
|---|---|
| T53.0X4* | Toxic effect of carbon tetrachloride, undetermined |
| T53.1X2* | Toxic effect of chloroform, intentional self-harm |
| T53.1X4* | Toxic effect of chloroform, undetermined |
| T53.2X2* | Toxic effect of trichloroethylene, intentional self-harm |
| T53.2X4* | Toxic effect of trichloroethylene, undetermined |
| T53.3X2* | Toxic effect of tetrachloroethylene, intentional self-harm |
| T53.3X4* | Toxic effect of tetrachloroethylene, undetermined |
| T53.4X2* | Toxic effect of dichloromethane, intentional self-harm |
| T53.4X4* | Toxic effect of dichloromethane, undetermined |
| T53.5X2* | Toxic effect of chlorofluorocarbons, intentional self-harm |
| T53.5X4* | Toxic effect of chlorofluorocarbons, undetermined |
| T53.6X2* | Toxic effect of other halogen derivatives of aliphatic hydrocarbons, intentional self-harm |
| T53.6X4* | Toxic effect of other halogen derivatives of aliphatic hydrocarbons, undetermined |
| T53.7X2* | Toxic effect of other halogen derivatives of aromatic hydrocarbons, intentional self-harm |
| T53.7X4* | Toxic effect of other halogen derivatives of aromatic hydrocarbons, undetermined |
| T53.92* | Toxic effect of unspecified halogen derivatives of aliphatic and aromatic hydrocarbons, intentional self-harm |
| T53.94* | Toxic effect of unspecified halogen derivatives of aliphatic and aromatic hydrocarbons, undetermined |
| T54.0X2* | Toxic effect of phenol and phenol homologues, intentional self-harm |
| T54.0X4* | Toxic effect of phenol and phenol homologues, undetermined |
| T54.1X2* | Toxic effect of other corrosive organic compounds, intentional self-harm |
| T54.1X4* | Toxic effect of other corrosive organic compounds, undetermined |
| T54.2X2* | Toxic effect of corrosive acids and acid-like substances, intentional self-harm |
| T54.2X4* | Toxic effect of corrosive acids and acid-like substances, undetermined |
| T54.3X2* | Toxic effect of corrosive alkalis and alkali-like substances, intentional self-harm |
| T54.3X4* | Toxic effect of corrosive alkalis and alkali-like substances, undetermined |
| T54.92* | Toxic effect of unspecified corrosive substance, intentional self-harm |
| T54.94* | Toxic effect of unspecified corrosive substance, undetermined |
| T55.0X2* | Toxic effect of soaps, intentional self-harm |
| T55.0X4* | Toxic effect of soaps, undetermined |
| T55.1X2* | Toxic effect of detergents, intentional self-harm |
| T55.1X4* | Toxic effect of detergents, undetermined |
| T56.0X2* | Toxic effect of lead and its compounds, intentional self-harm |
| T56.0X4* | Toxic effect of lead and its compounds, undetermined |
| T56.1X2* | Toxic effect of mercury and its compounds, intentional self-harm |
| T56.1X4* | Toxic effect of mercury and its compounds, undetermined |
| T56.2X2* | Toxic effect of chromium and its compounds, intentional self-harm |
| T56.2X4* | Toxic effect of chromium and its compounds, undetermined |
| T56.3X2* | Toxic effect of cadmium and its compounds, intentional self-harm |
| T56.3X4* | Toxic effect of cadmium and its compounds, undetermined |
| T56.4X2* | Toxic effect of copper and its compounds, intentional self-harm |
| T56.4X4* | Toxic effect of copper and its compounds, undetermined |
| T56.5X2* | Toxic effect of zinc and its compounds, intentional self-harm |
| T56.5X4* | Toxic effect of zinc and its compounds, undetermined |
| T56.6X2* | Toxic effect of tin and its compounds, intentional self-harm |
| T56.6X4* | Toxic effect of tin and its compounds, undetermined |
| T56.7X2* | Toxic effect of beryllium and its compounds, intentional self-harm |
| T56.7X4* | Toxic effect of beryllium and its compounds, undetermined |
| T56.812* | Toxic effect of thallium, intentional self-harm |
| T56.814* | Toxic effect of thallium, undetermined |
| T56.892* | Toxic effect of other metals, intentional self-harm |
| T56.894* | Toxic effect of other metals, undetermined |

| | |
|---|---|
| T56.8X2* | Toxic effect of other metals, intentional self-harm |
| T56.8X4* | Toxic effect of other metals, undetermined |
| T56.92* | Toxic effect of unspecified metal, intentional self-harm |
| T56.94* | Toxic effect of unspecified metal, undetermined |
| T57.0X2* | Toxic effect of arsenic and its compounds, intentional self-harm |
| T57.0X4* | Toxic effect of arsenic and its compounds, undetermined |
| T57.1X2* | Toxic effect of phosphorus and its compounds, intentional self-harm |
| T57.1X4* | Toxic effect of phosphorus and its compounds, undetermined |
| T57.2X2* | Toxic effect of manganese and its compounds, intentional self-harm |
| T57.2X4* | Toxic effect of manganese and its compounds, undetermined |
| T57.3X2* | Toxic effect of hydrogen cyanide, intentional self-harm |
| T57.3X4* | Toxic effect of hydrogen cyanide, undetermined |
| T57.8X2* | Toxic effect of other specified inorganic substances, intentional self-harm |
| T57.8X4* | Toxic effect of other specified inorganic substances, undetermined |
| T57.92* | Toxic effect of unspecified inorganic substance, intentional self-harm |
| T57.94* | Toxic effect of unspecified inorganic substance, undetermined |
| T58.02* | Toxic effect of carbon monoxide from motor vehicle exhaust, intentional self-harm |
| T58.04* | Toxic effect of carbon monoxide from motor vehicle exhaust, undetermined |
| T58.12* | Toxic effect of carbon monoxide from utility gas, intentional self-harm |
| T58.14* | Toxic effect of carbon monoxide from utility gas, undetermined |
| T58.2X2* | Toxic effect of carbon monoxide from incomplete combustion of other domestic fuels, intentional self-harm |
| T58.2X4* | Toxic effect of carbon monoxide from incomplete combustion of other domestic fuels, undetermined |
| T58.8X2* | Toxic effect of carbon monoxide from other source, intentional self-harm |
| T58.8X4* | Toxic effect of carbon monoxide from other source, undetermined |
| T58.92* | Toxic effect of carbon monoxide from unspecified source, intentional self-harm |
| T58.94* | Toxic effect of carbon monoxide from unspecified source, undetermined |
| T59.0X2* | Toxic effect of nitrogen oxides, intentional self-harm |
| T59.0X4* | Toxic effect of nitrogen oxides, undetermined |
| T59.1X2* | Toxic effect of sulfur dioxide, intentional self-harm |
| T59.1X4* | Toxic effect of sulfur dioxide, undetermined |
| T59.2X2* | Toxic effect of formaldehyde, intentional self-harm |
| T59.2X4* | Toxic effect of formaldehyde, undetermined |
| T59.3X2* | Toxic effect of lacrimogenic gas, intentional self-harm |
| T59.3X4* | Toxic effect of lacrimogenic gas, undetermined |
| T59.4X2* | Toxic effect of chlorine gas, intentional self-harm |
| T59.4X4* | Toxic effect of chlorine gas, undetermined |
| T59.5X2* | Toxic effect of fluorine gas and hydrogen fluoride, intentional self-harm |
| T59.5X4* | Toxic effect of fluorine gas and hydrogen fluoride, undetermined |
| T59.6X2* | Toxic effect of hydrogen sulfide, intentional self-harm |
| T59.6X4* | Toxic effect of hydrogen sulfide, undetermined |
| T59.7X2* | Toxic effect of carbon dioxide, intentional self-harm |
| T59.7X4* | Toxic effect of carbon dioxide, undetermined |
| T59.812* | Toxic effect of smoke, intentional self-harm |
| T59.814* | Toxic effect of smoke, undetermined |
| T59.892* | Toxic effect of other specified gases, fumes and vapors, intentional self-harm |
| T59.894* | Toxic effect of other specified gases, fumes and vapors, undetermined |
| T59.92* | Toxic effect of unspecified gases, fumes and vapors, intentional self-harm |
| T59.94* | Toxic effect of unspecified gases, fumes and vapors, undetermined |
| T60.0X2* | Toxic effect of organophosphate and carbamate insecticides, intentional self-harm |
| T60.0X4* | Toxic effect of organophosphate and carbamate insecticides, undetermined |
| T60.1X2* | Toxic effect of halogenated insecticides, intentional self-harm |

| | |
|---|---|
| T60.1X4* | Toxic effect of halogenated insecticides, undetermined |
| T60.2X2* | Toxic effect of other insecticides, intentional self-harm |
| T60.2X4* | Toxic effect of other insecticides, undetermined |
| T60.3X2* | Toxic effect of herbicides and fungicides, intentional self-harm |
| T60.3X4* | Toxic effect of herbicides and fungicides, undetermined |
| T60.4X2* | Toxic effect of rodenticides, intentional self-harm |
| T60.4X4* | Toxic effect of rodenticides, undetermined |
| T60.8X2* | Toxic effect of other pesticides, intentional self-harm |
| T60.8X4* | Toxic effect of other pesticides, undetermined |
| T60.92* | Toxic effect of unspecified pesticide, intentional self-harm |
| T60.94* | Toxic effect of unspecified pesticide, undetermined |
| T61.02* | Ciguatera fish poisoning, intentional self-harm |
| T61.04* | Ciguatera fish poisoning, undetermined |
| T61.12* | Scombroid fish poisoning, intentional self-harm |
| T61.14* | Scombroid fish poisoning, undetermined |
| T61.772* | Other fish poisoning, intentional self-harm |
| T61.774* | Other fish poisoning, undetermined |
| T61.782* | Other shellfish poisoning, intentional self-harm |
| T61.784* | Other shellfish poisoning, undetermined |
| T61.8X2* | Toxic effect of other seafood, intentional self-harm |
| T61.8X4* | Toxic effect of other seafood, undetermined |
| T61.92* | Toxic effect of unspecified seafood, intentional self-harm |
| T61.94* | Toxic effect of unspecified seafood, undetermined |
| T62.0X2* | Toxic effect of ingested mushrooms, intentional self-harm |
| T62.0X4* | Toxic effect of ingested mushrooms, undetermined |
| T62.1X2* | Toxic effect of ingested berries, intentional self-harm |
| T62.1X4* | Toxic effect of ingested berries, undetermined |
| T62.2X2* | Toxic effect of other ingested (parts of) plant(s), intentional self-harm |
| T62.2X4* | Toxic effect of other ingested (parts of) plant(s), undetermined |
| T62.8X2* | Toxic effect of other specified noxious substances eaten as food, intentional self-harm |
| T62.8X4* | Toxic effect of other specified noxious substances eaten as food, undetermined |
| T62.92* | Toxic effect of unspecified noxious substance eaten as food, intentional self-harm |
| T62.94* | Toxic effect of unspecified noxious substance eaten as food, undetermined |
| T63.002* | Toxic effect of unspecified snake venom, intentional self-harm |
| T63.004* | Toxic effect of unspecified snake venom, undetermined |
| T63.012* | Toxic effect of rattlesnake venom, intentional self-harm |
| T63.014* | Toxic effect of rattlesnake venom, undetermined |
| T63.022* | Toxic effect of coral snake venom, intentional self-harm |
| T63.024* | Toxic effect of coral snake venom, undetermined |
| T63.032* | Toxic effect of taipan venom, intentional self-harm |
| T63.034* | Toxic effect of taipan venom, undetermined |
| T63.042* | Toxic effect of cobra venom, intentional self-harm |
| T63.044* | Toxic effect of cobra venom, undetermined |
| T63.062* | Toxic effect of venom of other North and South American snake, intentional self-harm |
| T63.064* | Toxic effect of venom of other North and South American snake, undetermined |
| T63.072* | Toxic effect of venom of other Australian snake, intentional self-harm |
| T63.074* | Toxic effect of venom of other Australian snake, undetermined |
| T63.082* | Toxic effect of venom of other African and Asian snake, intentional self-harm |
| T63.084* | Toxic effect of venom of other African and Asian snake, undetermined |
| T63.092* | Toxic effect of venom of other snake, intentional self-harm |
| T63.094* | Toxic effect of venom of other snake, undetermined |
| T63.112* | Toxic effect of venom of gila monster, intentional self-harm |
| T63.114* | Toxic effect of venom of gila monster, undetermined |

| | |
|---|---|
| T63.122* | Toxic effect of venom of other venomous lizard, intentional self-harm |
| T63.124* | Toxic effect of venom of other venomous lizard, undetermined |
| T63.192* | Toxic effect of venom of other reptiles, intentional self-harm |
| T63.194* | Toxic effect of venom of other reptiles, undetermined |
| T63.2X2* | Toxic effect of venom of scorpion, intentional self-harm |
| T63.2X4* | Toxic effect of venom of scorpion, undetermined |
| T63.302* | Toxic effect of unspecified spider venom, intentional self-harm |
| T63.304* | Toxic effect of unspecified spider venom, undetermined |
| T63.312* | Toxic effect of venom of black widow spider, intentional self-harm |
| T63.314* | Toxic effect of venom of black widow spider, undetermined |
| T63.322* | Toxic effect of venom of tarantula, intentional self-harm |
| T63.324* | Toxic effect of venom of tarantula, undetermined |
| T63.332* | Toxic effect of venom of brown recluse spider, intentional self-harm |
| T63.334* | Toxic effect of venom of brown recluse spider, undetermined |
| T63.392* | Toxic effect of venom of other spider, intentional self-harm |
| T63.394* | Toxic effect of venom of other spider, undetermined |
| T63.412* | Toxic effect of venom of centipedes and venomous millipedes, intentional self-harm |
| T63.414* | Toxic effect of venom of centipedes and venomous millipedes, undetermined |
| T63.422* | Toxic effect of venom of ants, intentional self-harm |
| T63.424* | Toxic effect of venom of ants, undetermined |
| T63.432* | Toxic effect of venom of caterpillars, intentional self-harm |
| T63.434* | Toxic effect of venom of caterpillars, undetermined |
| T63.442* | Toxic effect of venom of bees, intentional self-harm |
| T63.444* | Toxic effect of venom of bees, undetermined |
| T63.452* | Toxic effect of venom of hornets, intentional self-harm |
| T63.454* | Toxic effect of venom of hornets, undetermined |
| T63.462* | Toxic effect of venom of wasps, intentional self-harm |
| T63.464* | Toxic effect of venom of wasps, undetermined |
| T63.482* | Toxic effect of venom of other arthropod, intentional self-harm |
| T63.484* | Toxic effect of venom of other arthropod, undetermined |
| T63.512* | Toxic effect of contact with stingray, intentional self-harm |
| T63.514* | Toxic effect of contact with stingray, undetermined |
| T63.592* | Toxic effect of contact with other venomous fish, intentional self-harm |
| T63.594* | Toxic effect of contact with other venomous fish, undetermined |
| T63.612* | Toxic effect of contact with Portugese Man-o-war, intentional self-harm |
| T63.614* | Toxic effect of contact with Portugese Man-o-war, undetermined |
| T63.622* | Toxic effect of contact with other jellyfish, intentional self-harm |
| T63.624* | Toxic effect of contact with other jellyfish, undetermined |
| T63.632* | Toxic effect of contact with sea anemone, intentional self-harm |
| T63.634* | Toxic effect of contact with sea anemone, undetermined |
| T63.692* | Toxic effect of contact with other venomous marine animals, intentional self-harm |
| T63.694* | Toxic effect of contact with other venomous marine animals, undetermined |
| T63.712* | Toxic effect of contact with venomous marine plant, intentional self-harm |
| T63.714* | Toxic effect of contact with venomous marine plant, undetermined |
| T63.792* | Toxic effect of contact with other venomous plant, intentional self-harm |
| T63.794* | Toxic effect of contact with other venomous plant, undetermined |
| T63.812* | Toxic effect of contact with venomous frog, intentional self-harm |
| T63.814* | Toxic effect of contact with venomous frog, undetermined |
| T63.822* | Toxic effect of contact with venomous toad, intentional self-harm |
| T63.824* | Toxic effect of contact with venomous toad, undetermined |
| T63.832* | Toxic effect of contact with other venomous amphibian, intentional self-harm |
| T63.834* | Toxic effect of contact with other venomous amphibian, undetermined |
| T63.892* | Toxic effect of contact with other venomous animals, intentional self-harm |

| | |
|---|---|
| T63.894* | Toxic effect of contact with other venomous animals, undetermined |
| T63.92* | Toxic effect of contact with unspecified venomous animal, intentional self-harm |
| T63.94* | Toxic effect of contact with unspecified venomous animal, undetermined |
| T64.02* | Toxic effect of aflatoxin, intentional self-harm |
| T64.04* | Toxic effect of aflatoxin, undetermined |
| T64.82* | Toxic effect of other mycotoxin food contaminants, intentional self-harm |
| T64.84* | Toxic effect of other mycotoxin food contaminants, undetermined |
| T65.0X2* | Toxic effect of cyanides, intentional self-harm |
| T65.0X4* | Toxic effect of cyanides, undetermined |
| T65.1X2* | Toxic effect of strychnine and its salts, intentional self-harm |
| T65.1X4* | Toxic effect of strychnine and its salts, undetermined |
| T65.212* | Toxic effect of chewing tobacco, intentional self-harm |
| T65.214* | Toxic effect of chewing tobacco, undetermined |
| T65.222* | Toxic effect of tobacco cigarettes, intentional self-harm |
| T65.224* | Toxic effect of tobacco cigarettes, undetermined |
| T65.292* | Toxic effect of other tobacco and nicotine, intentional self-harm |
| T65.294* | Toxic effect of other tobacco and nicotine, undetermined |
| T65.3X2* | Toxic effect of nitroderivatives and aminoderivatives of benzene and its homologues, intentional self-harm |
| T65.3X4* | Toxic effect of nitroderivatives and aminoderivatives of benzene and its homologues, undetermined |
| T65.4X2* | Toxic effect of carbon disulfide, intentional self-harm |
| T65.4X4* | Toxic effect of carbon disulfide, undetermined |
| T65.5X2* | Toxic effect of nitroglycerin and other nitric acids and esters, intentional self-harm |
| T65.5X4* | Toxic effect of nitroglycerin and other nitric acids and esters, undetermined |
| T65.6X2* | Toxic effect of paints and dyes, not elsewhere classified, intentional self-harm |
| T65.6X4* | Toxic effect of paints and dyes, not elsewhere classified, undetermined |
| T65.812* | Toxic effect of latex, intentional self-harm |
| T65.814* | Toxic effect of latex, undetermined |
| T65.822* | Toxic effect of harmful algae and algae toxins, intentional self-harm |
| T65.824* | Toxic effect of harmful algae and algae toxins, undetermined |
| T65.832* | Toxic effect of fiberglass, intentional self-harm |
| T65.834* | Toxic effect of fiberglass, undetermined |
| T65.892* | Toxic effect of other specified substances, intentional self-harm |
| T65.894* | Toxic effect of other specified substances, undetermined |
| T65.92* | Toxic effect of unspecified substance, intentional self-harm |
| T65.94* | Toxic effect of unspecified substance, undetermined |
| T71.112* | Asphyxiation due to smothering under pillow, intentional self-harm |
| T71.114* | Asphyxiation due to smothering under pillow, undetermined |
| T71.122* | Asphyxiation due to plastic bag, intentional self-harm |
| T71.124* | Asphyxiation due to plastic bag, undetermined |
| T71.132* | Asphyxiation due to being trapped in bed linens, intentional self-harm |
| T71.134* | Asphyxiation due to being trapped in bed linens, undetermined |
| T71.144* | Asphyxiation due to smothering under another person's body (in bed), undetermined |
| T71.152* | Asphyxiation due to smothering in furniture, intentional self-harm |
| T71.154* | Asphyxiation due to smothering in furniture, undetermined |
| T71.162* | Asphyxiation due to hanging, intentional self-harm |
| T71.164* | Asphyxiation due to hanging, undetermined |
| T71.192* | Asphyxiation due to mechanical threat to breathing due to other causes, intentional self-harm |
| T71.194* | Asphyxiation due to mechanical threat to breathing due to other causes, undetermined |
| T71.222* | Asphyxiation due to being trapped in a car trunk, intentional self-harm |
| T71.224* | Asphyxiation due to being trapped in a car trunk, undetermined |
| T71.232* | Asphyxiation due to being trapped in a (discarded) refrigerator, intentional self-harm |
| T71.234* | Asphyxiation due to being trapped in a (discarded) refrigerator, undetermined |

| | |
|---|---|
| X71* | Intentional self-harm by drowning and submersion |
| X72* | Intentional self-harm by handgun discharge |
| X73* | Intentional self-harm by rifle, shotgun and larger firearm discharge |
| X74* | Intentional self-harm by other and unspecified firearm and gun discharge |
| X75* | Intentional self-harm by explosive material |
| X76* | Intentional self-harm by smoke, fire and flames |
| X77* | Intentional self-harm by steam, hot vapors and hot objects |
| X78* | Intentional self-harm by sharp object |
| X79* | Intentional self-harm by blunt object |
| X80* | Intentional self-harm by jumping from a high place |
| X81* | Intentional self-harm by jumping or lying in front of moving object |
| X82* | Intentional self-harm by crashing of motor vehicle |
| X83* | Intentional self-harm by other specified means |
| Y21* | Drowning and submersion, undetermined intent |
| Y22* | Handgun discharge, undetermined intent |
| Y23* | Rifle, shotgun and larger firearm discharge, undetermined intent |
| Y24* | Other and unspecified firearm discharge, undetermined intent |
| Y25* | Contact with explosive material, undetermined intent |
| Y26* | Exposure to smoke, fire and flames, undetermined intent |
| Y27* | Contact with steam, hot vapors and hot objects, undetermined intent |
| Y28* | Contact with sharp object, undetermined intent |
| Y29* | Contact with blunt object, undetermined intent |
| Y30* | Falling, jumping or pushed from a high place, undetermined intent |
| Y31* | Falling, lying or running before or into moving object, undetermined intent |
| Y32* | Crashing of motor vehicle, undetermined intent |
| Y33* | Other specified events, undetermined intent |
| Z91.5 | Personal history of self-harm |

**Table S5** Top 50 keywords generated by word2vec as semantically similar to 'suicide' and 'suicidal'.

| | suicide+ suicidal | | suicide | | suicidal | |
|---|---|---|---|---|---|---|
| | context size = 5 | context size = 15 | context size = 5 | context size = 15 | context size = 5 | context size = 15 |
| 1 | suicide | suicide | self-harm | manic | ideation | ideation |
| 2 | suicidal | suicidal | suicidal | ideation | homicidal | homicidal |
| 3 | ideation | ideation | paranoid | suicidal | ideations | ideations |
| 4 | homicidal | homicidal | homicide | self-harm | paranoia | suicidality |
| 5 | self-harm | ideations | ideation | suicided | suidical | paranoia |
| 6 | ideations | manic | suicide/homicide | mania | suicidality | suidical |
| 7 | paranoia | self-harm | self-mutilation | homicidal | self-harm | delusional |
| 8 | paranoid | mania | paranoia | s/h | delusional | self-harm |
| 9 | suidical | suicidality | self-harm--plan | self-mutilation | paranoid | thoughts |
| 10 | suicidality | paranoia | manic | ptsd | suicidial | mania |
| 11 | manic | suidical | mutilation | paranoid | homocidal | manic |
| 12 | self-mutilation | self-mutilation | schizophrenic | mutilation | si/hi | si/hi |
| 13 | delusional | hypomanic | suicided | ideations | homicidal/suicidal | hypomanic |
| 14 | homicial | delusional | codependent | threats | homicial | homocidal |
| 15 | mutilation | paranoid | w/bipolar | codependent | sucidal | self-mutilation |
| 16 | sucidal | s/h | homicidal | suidical | intent/plan | suicide |
| 17 | suicidial | homocidal | self-harm, | self-destructive | avh | overdosing |
| 18 | parasuicidal | self-destructive | homicial | paranoia | ideation/plan | homicidality |
| 19 | intent/plan | mutilation | ideations | hypomanic | thoughts | paranoid |
| 20 | si/hi | si/hi | parasuicidal | parasuicidal | si/hi/intent | grandiose |
| 21 | overdosing | parasuicidal | houghts | strife | suicide | self-destructive |
| 22 | hypomanic | overdosing | then-current | flashbacks | manic | avh |
| 23 | then-current | psychotropic | overdosing | suicidality | =+@si | grandeur |
| 24 | avh | flashbacks | none--intent | none--intent | ;{deny | persecutory |
| 25 | si/hi/intent | persecutory | schizophrenia | psychotropic | urge/intent | s/h |
| 26 | homicidal/suicidal | persecution | overdoses | bpad | persecution | mutilation |

| | | | | | | |
|---|---|---|---|---|---|---|
| 27 | psychiatrically | threats | suidical | dwf | self-mutilation | ruminations |
| 28 | ideation/plan | isolative | hypomanic | "manic | hypomanic | parasuicidal |
| 29 | =+@si | thoughts | ptsd | isolative | =+@suicidal | suicidial |
| 30 | mania/hypomania | manic/hypomanic | delusional | i/j | psychiatrically | manic/hypomanic |
| 31 | homocidal | grandiose | mania/hypomania | j/i | suicidal/homicidal | future-oriented |
| 32 | homicide | helplessness | rapes | homocidal | ideation{ | persecution |
| 33 | self-destructive | mania/hypomania | threats | homicide | overdosing | psychotic |
| 34 | suicidal/homicidal | suicided | s/h | plan/intent | parasuicidal | "depressed |
| 35 | mania | ptsd | overdose | persecutory | grandeur | psychotropic |
| 36 | urge/intent | avh | psychiatrically | overdosing | persecutory | hopelessness |
| 37 | plan/intent | plan/intent | rape | suicidal/homicidal | plan/intent | helplessness |
| 38 | persecution | "hopeless | saam | homosexuality | grandiose | illusions |
| 39 | self-harm--plan | psychotic | sucidal | reckless | mania/hypomania | isolative |
| 40 | thoughts | homicidality | self-destructive | manic/hypomanic | then-current | mood-congruent |
| 41 | s/h | fidgetiness | "schizophrenia | mania/hypomania | manic/hypomanic | flashbacks |
| 42 | psychotropic | future-oriented | suicidality | nihilistic | mutilation | mania/hypomania |
| 43 | plans/intent | suicidal/homicidal | bpad | helplessness | delusions | obsessions |
| 44 | persecutory | "manic | intent/plan | hypervigilence | hopelessness | urge/intent |
| 45 | threats | hypervigilance | mania | fidgetiness | plans/intent | "hopeless |
| 46 | grandeur | ruminations | si/hi/intent | estrangement | preoccupations | pressured |
| 47 | suicidal/ | hopelessness | murder | "hopeless | mania | fidgetiness |
| 48 | ;{deny | obsessions | psychotropic | discord | ideation/intent/plan | worthless |
| 49 | overdoses | suicidial | suicide, | =--@plan | suicideal | dysphoric |
| 50 | self-harm, | hopless | avh | self-harm--plan-no-intent-no--past | ruminations | hypervigilance |

**Table S6** Query terms used to retrieve suicidal ideation and suicide attempt.

| Phenotype | Query terms |
|---|---|
| Suicidal Ideation | suicid(al\|e) idea(tion\|s)<br>suicid(al\|e) thought(s)<br>thought(s) of suicide<br>want(s\|ing) to die<br>(thoughts\|think\|want)(s\|ing) (of\|to) (take\|end)(ing) (my\|his\|her) (own) life<br>(thoughts\|think\|want)(s\|ing) (of\|to) (kill\|shoot\|hang\|poison)(ing) (myself\|himself\|herself)<br>feel(s\|ing) (very) suicidal |
| Suicide attempt | suicid(al\|e) attempt<br>(try\|tried\|tries\|attempted\|attempts) (of\|to) (take\|end)(ing) (my\|his\|her) (own) life<br>(try\|tried\|tries\|attempted\|attempts) (of\|to) (kill\|shoot\|hang\|poison)(ing) (myself\|himself\|herself) |

**Table S7** Comparative analysis for extracting the top K highest ranked suicidal ideation (SI) and suicide attempt (SA) patients using various configurations of the suicide label assignment method. The "All retrieved" experiments include all the patients retrieved by the NLP system while the "w/ 1+ positive" ones use only patients with at least one positive suicide mention in their notes. "NLP" and "NLP+ICD" experiments are associated with methods using $p_{rank}$ and $p_{NLP+ICD}$, respectively.

| | | All retrieved | | | | w/ 1+ positive | | | |
|---|---|---|---|---|---|---|---|---|---|
| | | NLP | | NLP+ICD | | NLP | | NLP+ICD | |
| | | mean | sd | mean | sd | mean | sd | mean | sd |
| SI | Top K (P@K=90) | 926.83 | 104.83 | 1,162.55 | 80.61 | 975.01 | 106.22 | 1,228.01 | 84.03 |
| | Top K (P@K=80) | 3,094.51 | 144.54 | 5,782.72 | 192.52 | 3,135.62 | 138.99 | 5,945.07 | 186.15 |
| SA | Top K (P@K=90) | 384.52 | 32.27 | 495.58 | 55.82 | 394.59 | 37.89 | 526.77 | 67.84 |
| | Top K (P@K=80) | 818.00 | 64.90 | 1,663.73 | 114.22 | 839.86 | 66.88 | 1,693.19 | 122.55 |

**Table S8** Suicidal ideation (SI) and suicide attempt (SA) cases extracted from: 1) the top ranked patients by the NLP system, 2) patients manually labeled as cases, 3) patients with positive assertions for suicidal ideation and suicide attempt in their psychiatric forms, and 4) patients with ICD10CM codes for self-injurious thoughts and behaviors.

| Resource | SI | SA |
|---|---:|---:|
| NLP, top K, P@K=90 | 1,209 | 380 |
| NLP, top K, P@K=80 | 5,342 | 1,384 |
| Chart review (cases) | 991 | 718 |
| Psychiatric forms (cases) | 5,232 | 4,275 |
| ICD10CM codes | 20,037 | 15,729 |
